# Modeling the “Bomb-Like” Dynamics of COVID-19 with Undetected Transmissions and the Implications for Policy

**DOI:** 10.1101/2020.04.05.20054338

**Authors:** Gary Lin, Anindya Bhaduri, Alexandra T. Strauss, Maxwell Pinz, Diego A. Martinez, Katie K. Tseng, Oliver Gatalo, Andrew T. Gaynor, Efrain Hernandez-Rivera, Emily Schueller, Yupeng Yang, Simon A. Levin, Eili Y. Klein, For the CDC MInD-Healthcare Program

## Abstract

Understanding the transmission dynamics of the severe acute respiratory syndrome coronavirus 2 (SARS-COV-2) is critical to inform sound policy decisions. We demonstrate how the transmission of undetected cases with pre-symptomatic, asymptomatic and mild symptoms, which are typically underreported due to lower testing capacity, explained the “bomb-like” behavior of exponential growth in the coronavirus disease 2019 (COVID-19) cases during early stages before the effects of large-scale, non-pharmaceutical interventions such as social distancing, school closures, or lockdowns. Using a Bayesian approach to epidemiological compartmental modeling, we captured the initial stages of the pandemic resulting in the explosion of cases and compared the parameter estimation with empirically measured values from the current knowledgebase. Parameter estimation was conducted using Markov chain Monte Carlo (MCMC) sampling methods with a Bayesian inference framework to estimate the proportion of undetected cases. Using data from the exponential phase of the pandemic prior to the implementation of interventions we estimated the basic reproductive number (*R*_*0*_) and symptomatic rates in Italy, Spain, South Korea, New York City, and Chicago. From this modeling study, *R*_*0*_ was estimated to be 3·25 (95% CrI, 1·09-29·77), 3·62 (95% CrI, 1·13-34·89), 2·75 (95% CrI, 1·04-22·44), 3·31 (95% CrI, 1·69-20·55), and 3·46 (95% CrI, 1·01-34·41), respectively. For all locations, 3-25% of infected patients were identified with moderate and severe symptoms in the early stage of the COVID-19 pandemic. Our modeling results support the mounting evidence that potentially large fractions of the infected population were undetected with asymptomatic and mild symptoms. Furthermore, a significant number of models of transmission that do not account for these asymptomatic cases may lead to an underestimation of *R*_*0*_ and, subsequently, policies that do not sufficiently reduce transmission to contain the spread of the virus. Detecting asymptomatic transmission can help slow down the spread of SARS-CoV-2.

**Author Summary:** The spread of SARS-CoV-2 has led to a global pandemic that is still spreading across countries. We fitted a mathematical model to reported infections in Spain, Italy, South Korea, New York, and Chicago before any large-scale interventions, such as lockdowns and school closures, and found that undetected infected individuals drove the accelerated pace of transmissions. Due to the limited capacity in testing in many of the five locations, undetected cases were most likely asymptomatic and mild to moderate symptomatic infections. Given the explosive nature in the number of cases during the early phase of the pandemic and the latest serological surveys, our study suggested that most active cases were undetected. Other cohort studies have shown that a significant proportion of cases reported little or no symptoms. We also showed that early detection of asymptomatic and mild symptomatic cases can lead to a slower spread of SARS-CoV-2 as evident in South Korea. Policies targeting symptomatic individuals, such as travel restrictions on affected areas or quarantines of sick individuals, are not as effective because they neglect asymptomatic transmission events.

## Introduction

The spread of the severe acute respiratory syndrome coronavirus 2 (SARS-CoV-2) was declared a global pandemic by the World Health Organization on March 11, and confirmed cases of coronavirus disease 2019 (COVID-19) have since grown exponentially on every inhabited continent [1]. Despite the rising numbers of cases and deaths, vital questions about the dynamics of the spread of COVID-19, particularly the “bomb-like” dynamics of the disease, remain. In most countries, states, and cities, the number of confirmed cases remained low for weeks and then suddenly exploded, with the number of confirmed cases exponentially increasing in a matter of days (Figure 1). There are two possible explanations for this dynamic: (i) the growth rate is intrinsic to the disease or (ii) the rapid growth is due to undercounting of asymptomatic or mild cases of the disease. The *R*_*0*_ measure is important for capturing the magnitude and speed that the population will be infected with SARS-CoV-2. Initial estimates of *R*_*0*_ ranged widely from 1·4 to 7·1 [2–6]. The variability in reported *R*_*0*_ between locations could be explained by different detection rates of asymptomatic and mild symptomatic cases.

**Figure 1.**
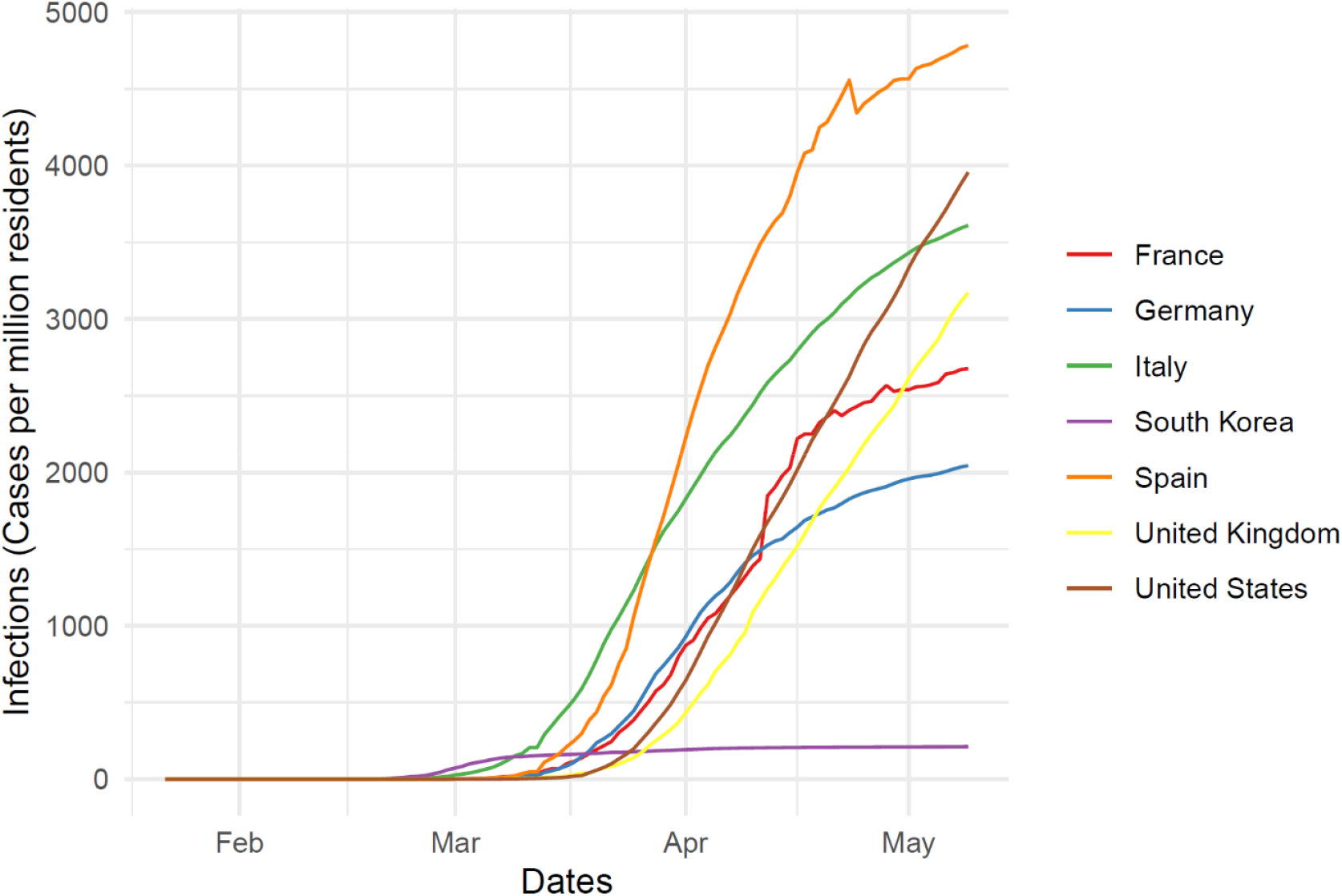
Confirmed cases of COVID-19 per million residents for France, Germany, Italy, South Korea, Spain, United Kingdon, and the United States. The rate of exponential increase are observed for all countries except South Korea where early surveillence was employed. Source: Center for Systems Science and Engineering [1] at Johns Hopkins Universty.

Understanding the percentage of the population infected with SARS-CoV-2 is critical for developing policies for containment, particularly as the threat of a second wave of disease appears inevitable. Most countries have instituted extended shutdowns to limit social contacts and opportunities for disease spread; the lifting of these restrictions in the continued presence of cases may expose these countries to explosive increases in infections. However, questions remain regarding the most effective strategies to contain the pandemic in the short- and long-term. Countries that have contained or slowed the spread of disease include China, Singapore, Hong Kong, Taiwan, and South Korea [7–11]. Through comprehensive strategies including early detection, extensive testing, shelter-in-place orders, and isolation of infected individuals, these countries have demonstrated that a combination of control measures can effectively slow transmission from affected individuals whether they are severely sick or asymptomatically infected. In contrast, isolated measures like broad travel restrictions or enforced quarantine have proven less effective, as the ability to achieve “zero risk” through these measures is virtually unattainable in most contexts [12,13]. Moreover, the effectiveness of these strategies, which primarily target individuals with noticeable symptoms of COVID-19, is questionable given evidence of asymptomatic and mildly symptomatic carriers as important vectors for community transmission of SARS-CoV-2 [14–17].

Emerging evidence from China [17], Germany [18], Taiwan [19], Iceland [20], and other places [21] suggests a larger fraction of the population may be asymptomatically or mildly infected than previously thought. The first report of genomic differences in the virus from Washington State in the United States suggested widespread community transmission as early as late January [22], which presumably generated asymptomatic or mild cases that were not identified. Given that the spread of the disease coincided with peak influenza season, it is reasonable to assume that mild COVID-19 infections could have been misdiagnosed and undetected. Furthermore, data on SARS-CoV-2’s effect on children have been almost entirely lacking in the numbers of cases and hospitalizations reported [23]. Some evidence suggests children are being infected at the same rate as adults, albeit with lesser severity [24,25], implying that at least 15% of the population in many countries may be asymptomatic when infected, with potential to be much higher. Studies have also emerged showing similar viral loads between asymptomatic and symptomatic cases, which suggests that there is potential for similar transmissibility between the two cohorts [18,26].

Here we assess the value of *R*_*0*_ using a Bayesian framework to estimate the role of asymptomatic cases in driving disease dynamics. Our goal was to explain the data associated with emerging outbreaks in countries where initial effects of distancing and other measures to control the disease were largely absent. During this early, “bomb-like” phase, infection count data are assumed to be largely representative of the transmission dynamics, with some proportion of the infected population being masked due to asymptomatic and mild symptomatic cases and shortages in testing. We examined data from Italy and Spain, both of which had large outbreaks that were not well contained and did not institute early lockdowns. These countries are compared to South Korea, which largely contained its outbreak through extensive testing and no hard lockdown. In addition, we contrasted these country-wide outbreaks with two metropolitan areas in the United States, New York City and Chicago, as dynamics at the city level may differ from those at the country level (see S1 Appendix for metropolitan area classification). Understanding the transmission patterns of SARS-CoV-2 can help policymakers predict critical moments in its progression, such as peak infection, the point when herd immunity has been reached, and local peaks during periods of relaxed or tightened restrictions on activity.

## Methods

We adapted the Kermack and McKendrick compartmental model to the reported disease dynamics of COVID-19 [27]. In our model, we assume there is (i) an incubation period for susceptible individuals that become infected; and (ii) a fraction of individuals who are asymptomatic or mildly symptomatic and neither tested nor counted as confirmed cases. For a given population *N*, the model is described by the following set of ordinary differential equations,

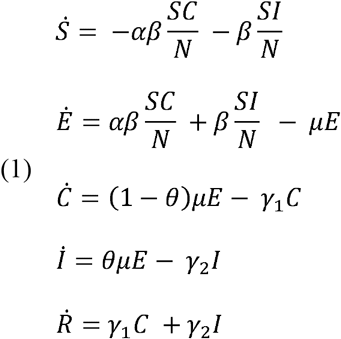

where *S* is the population of susceptible individuals, and *E* are exposed individuals incubating the disease that eventually become infected. After an incubation period, *μ*^*-1*^, we assume a proportion of individuals, defined by *θ*, will transition to a state *I* with moderate to severe symptoms which would result in detection, while the other proportion *C* will remain undetected because they have mild symptoms or are asymptomatic. For undetected individuals, we assumed there is a reduction of *α* which modifies the symptomatic transmission rate *β*. Finally, *R* represents individuals that either recover, die, or remove themselves from transmission through self-quarantine until they are no longer transmissible at rates γ_1_ for asymptomatic/mild cases and γ_2_ for symptomatic cases.

### Bayesian estimation of model parameters

While traditional unbiased curve-fitting methods yield a set of parameter estimations that capture observed data, they do not account for known prior belief on parameter ranges shown in Table 1. A Bayesian approach to parameter estimation allows us to quantify the credibility of one set of model parameters. This approach is useful for this context as it provides a range of credible parameters that describes the observed data and allows us to infer underlying causal pathways and quantify uncertainty.

**Table 1.**
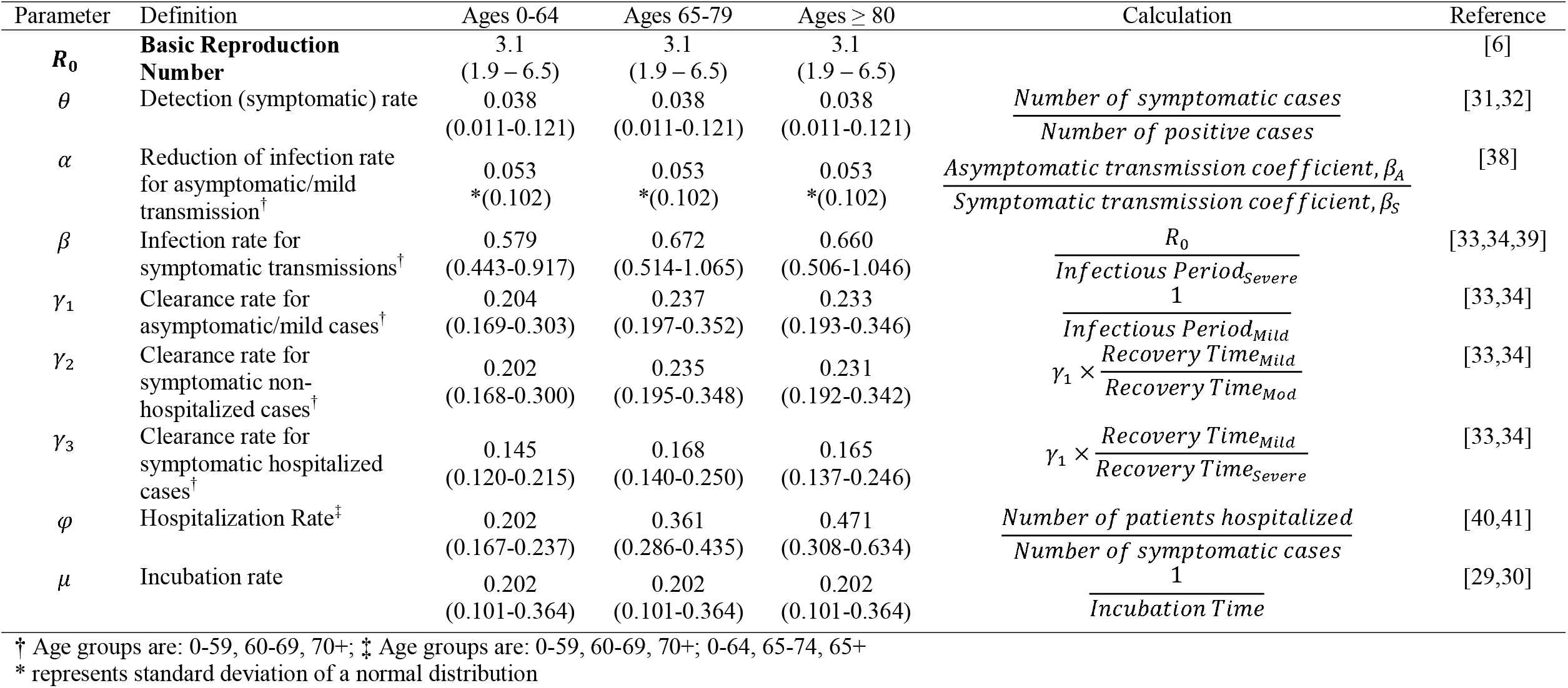
Parameters estimation from other literature where the means and corresponding credible intervals are shown in the parenthesis

To estimate the posterior distribution of the parameters, we obtained data on outbreaks in Italy, Spain, South Korea, and Chicago from the Center for Systems Science and Engineering at Johns Hopkins University [1], and for New York City we fitted the model to reported hospitalizations of COVID-19 patients obtained from the New York Department of Health and Mental Health [28]. We trained the model to fit the observed data assuming that the first observed case was in the more severe, symptomatic group. For each initial symptomatic case, we seeded an additional twenty people in the exposed compartment. We determined these timeframes in 2020 to be January 31 to March 21 for Italy, February 1 to March 25 for Spain, January 22 to March 3 for South Korea, January 22 to April 22 for Chicago, and February 17 to April 10 for New York City, which coincide with the first observed case.

We used Markov Chain Monte Carlo (MCMC) methods assuming a uniform prior density, specifically the Metropolis-Hasting Algorithm, to approximate a posterior distribution of the parameter set. To explore all possibilities and demonstrate robustness in the estimation results, we ran three cases for each locale with different Bayesian priors for the symptomatic rate (*θ*). Reported parameter estimates are the medians of the posterior distributions and the 95% credible intervals (CrI) from quantiles of the posterior distribution. We provide a full description of the mathematical structure of our models and estimation procedures in the S1 Appendix. The model was fit to the number of cumulative infections and the likelihood was estimated based on the sum of squared residuals (SSR). For New York City, we included the number of cumulative hospitalizations into the SSR error metric in order to estimate the hospitalization rate. The error between the results of the model yielding the expected number of observed cases and the observed number of cases is considered to be distributed with 𝒩(0, σ^2^).

### Biological Parameters and Prior Beliefs

In order to assess fit, determine model parameter priors, and provide context for the analysis, we conducted a literature search of the biology and transmission of SARS-CoV-2 using peer-reviewed literature and non-peer-reviewed literature available on pre-print servers medRxiv, bioRxiv and SSRN’s First Look (Table 1). From the literature, we found a mean incubation period, defined as the time from exposure to onset of illness, of 4·95 (CI, 2·75-9·90) [29,30]. The proportion of the population with no/mild/moderate symptoms was estimated to be as high as 96·2% of the population [31,32]. Based on the dynamics of earlier coronaviruses [33], the recovery periods for symptomatic and asymptomatic cases were estimated to be ∼4·9 days while hospitalized cases were around 6·9 days [33,34]. In our model, γ_1,_ γ_2,_ and γ_3_ represented the infectiousness period, which is not necessarily equivalent to the recovery rate since severe/moderate symptomatic and hospitalized cases are effectively removed due to self-quarantine. Data on confirmed cases reported in the US estimate the hospitalization rate for all ages to range from 20·7%-31·4%, with lower rates observed among younger populations and higher rates observed among older age groups. Finally, we estimated the transmission rates by asymptomatic, *αβ*, and symptomatic, *β*, persons based on calculations of the basic reproduction number, *R*_*0*_, of COVID-19, clearance estimates, and contact rates. Values for *R*_*0*_ in the literature range from 1·4 to 7·1 [2–6]; given these values we assumed the transmission rate for *β* was likely between 0·01 and 2.

We conducted MCMC sampling for three cases using different bounds on weakly-informed priors for the symptomatic rate in order to test for robustness. The main results are presented as the fitted parameter values of the posterior distribution with unconstrained symptomatic rate (*θ*) priors and their relative relationship to the known biology. In addition to the unconstrained prior of *θ*, we refitted the data twice using two different prior assumptions of the symptomatic fraction: *θ* is less than 50%, and *θ* is less than 10% with the results located in S1 Appendix. In our parameter estimation, we assumed that transmission from our unobserved group was lower by a factor of *α* than transmission from the symptomatic observed group, *β*, as mild/asymptomatic cases are likely to shed fewer viruses and thus be less transmissible. Thus, we constrained *α* between 0·01-0·99 and *β* between 0·01-2·00. As the process is stochastic, we evaluated the median and credible intervals of the posterior distribution obtained from this procedure to guide our understanding of the parameters and to assess the ability of the model to fit actual data with biologically plausible values.

## Results

Assuming a uniform prior with no constraint on the fraction of the population that was symptomatic, *θ*, the parameter estimation from our MCMC sampling resulted in a posterior median for the symptomatic rate of 0·04 (95% CrI, 0·01-0·41), 0·03 (95% CrI, 0·01-0·32), 0·18 (95% CrI, 0·02-0·85) for Italy, Spain, and South Korea, respectively, while the symptomatic rates of metropolitan areas of New York City and Chicago were 0·25 (95% CrI, 0·01-0·89) and 0·03 (95% CrI, 0·01-0·36), respectively. The median posterior values of these parameter ranges resulted in a calculated *R*_*0*_ of 3·25 (95% CrI, 1·09-29·77), 3·62 (95% CrI, 1·13-34·89), 2·75 (95% CrI, 1·04-22·44), 3·31 (95% CrI, 1·69-20·55), and 3·46 (95% CrI, 1·01-34·41) for Italy, Spain, South Korea, New York City, and Chicago, respectively (Table 2 and Figure 2).

**Table 2.**
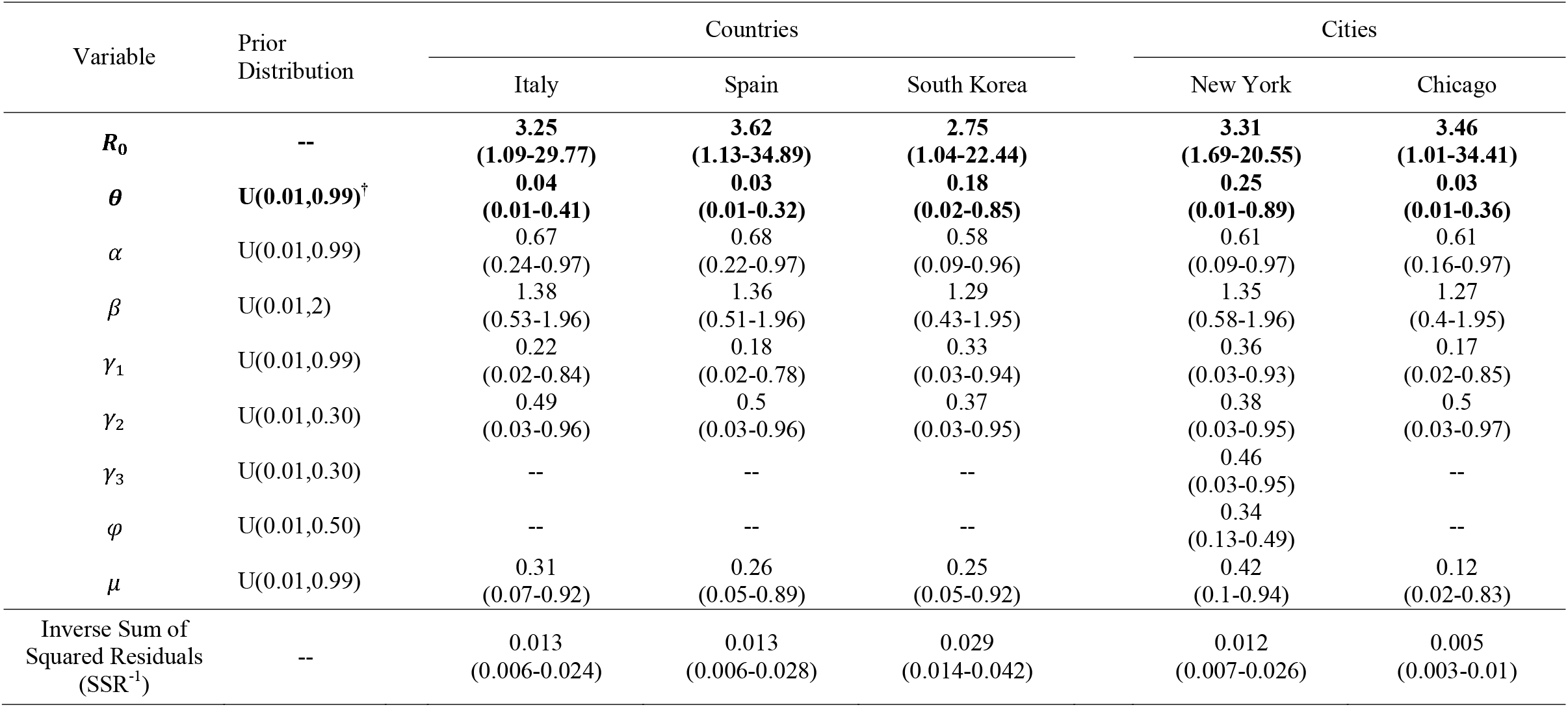
Posterior parameters estimation for US cities from the Bayesian analysis using the epidemiological model where the medians and corresponding 95% credible intervals are shown in the parenthesis.

| Variable | Prior Distribution | Countries |  |  | Cities |  |
| --- | --- | --- | --- | --- | --- | --- |
|  |  | Italy | Spain | South Korea | New York | Chicago |
| $R_0$ | -- | **3.25**<br>(1.09-29.77) | **3.62**<br>(1.13-34.89) | **2.75**<br>(1.04-22.44) | **3.31**<br>(1.69-20.55) | **3.46**<br>(1.01-34.41) |
| $\theta$ | $U(0.01, 0.99)^\dagger$ | **0.04**<br>(0.01-0.41) | **0.03**<br>(0.01-0.32) | **0.18**<br>(0.02-0.85) | **0.25**<br>(0.01-0.89) | **0.03**<br>(0.01-0.36) |
| $\alpha$ | $U(0.01, 0.99)$ | 0.67<br>(0.24-0.97) | 0.68<br>(0.22-0.97) | 0.58<br>(0.09-0.96) | 0.61<br>(0.09-0.97) | 0.61<br>(0.16-0.97) |
| $\beta$ | $U(0.01, 2)$ | 1.38<br>(0.53-1.96) | 1.36<br>(0.51-1.96) | 1.29<br>(0.43-1.95) | 1.35<br>(0.58-1.96) | 1.27<br>(0.4-1.95) |
| $\gamma_1$ | $U(0.01, 0.99)$ | 0.22<br>(0.02-0.84) | 0.18<br>(0.02-0.78) | 0.33<br>(0.03-0.94) | 0.36<br>(0.03-0.93) | 0.17<br>(0.02-0.85) |
| $\gamma_2$ | $U(0.01, 0.30)$ | 0.49<br>(0.03-0.96) | 0.5<br>(0.03-0.96) | 0.37<br>(0.03-0.95) | 0.38<br>(0.03-0.95) | 0.5<br>(0.03-0.97) |
| $\gamma_3$ | $U(0.01, 0.30)$ | -- | -- | -- | 0.46<br>(0.03-0.95) | -- |
| $\varphi$ | $U(0.01, 0.50)$ | -- | -- | -- | 0.34<br>(0.13-0.49) | -- |
| $\mu$ | $U(0.01, 0.99)$ | 0.31<br>(0.07-0.92) | 0.26<br>(0.05-0.89) | 0.25<br>(0.05-0.92) | 0.42<br>(0.1-0.94) | 0.12<br>(0.02-0.83) |
| Inverse Sum of Squared Residuals ($SSR^{-1}$) | -- | 0.013<br>(0.006-0.024) | 0.013<br>(0.006-0.028) | 0.029<br>(0.014-0.042) | 0.012<br>(0.007-0.026) | 0.005<br>(0.003-0.01) |

**Figure 2.**
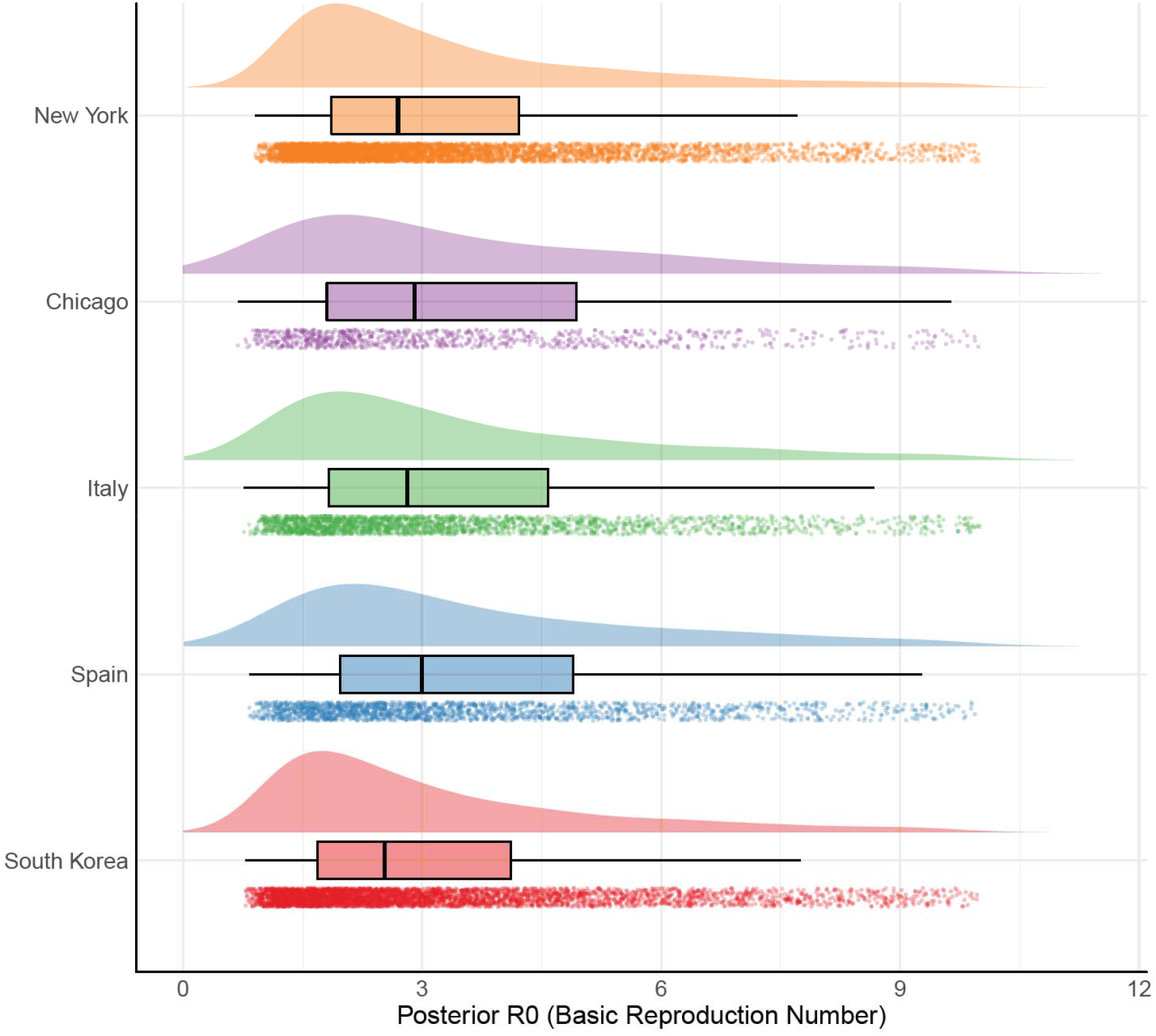
Estimated Basic Reproduction Number *R*_*0*_ for New York, Chicago, Italy, Spain, and South Korea from Bayesian distribution. Each curve is the posterior distribution generated from the MCMC sampling for the *R*_*0*_ of each location are shown. For each location, the raw data, box and whisker plot (median, interquartile ranges, 95% credible intervals), and probability density are displayed top-to-bottom.

Variation between the locales that had higher symptomatic infections was primarily governed by differences in the length of the infectiousness of the asymptomatic group. In the high symptomatic scenarios of South Korea and New York City, the fit of the parameter produced median infectious periods of 3 days in contrast to lower symptomatic locales where the infectious period was estimated to be about 5-6 days. The symptomatic group at all locales had median infectious periods of 2-3 days.

Assuming stronger priors with constrained uniform priors of the symptomatic rate *θ* to be under 50% or 10%, the inverse sum of squared residuals (SSR^-1^) was relatively lower, resulting in a slightly better fit for all locations (Tables S1 and S2 in S1 Appendix). However, the parameter estimation of constrained priors still resulted in similar values as to their unconstrained counterparts. The posterior density of *θ* was truncated for South Korea and New York City if we constrained *θ*’s prior to be less than 10 percent (Tables S31 and S32 in S1 Appendix). Figure 3 illustrates a negative relationship between symptomatic rate *θ* and SSR^-1^, indicating that lower estimates of symptomatic rates resulted in higher posterior likelihoods. In all projections using the accepted posterior parameter samples, the simulations overestimated actual case numbers following the initial period of infection, as non-pharmaceutical interventions such as lockdowns artificially reduced disease spread (Figure 4).

**Figure 3.**
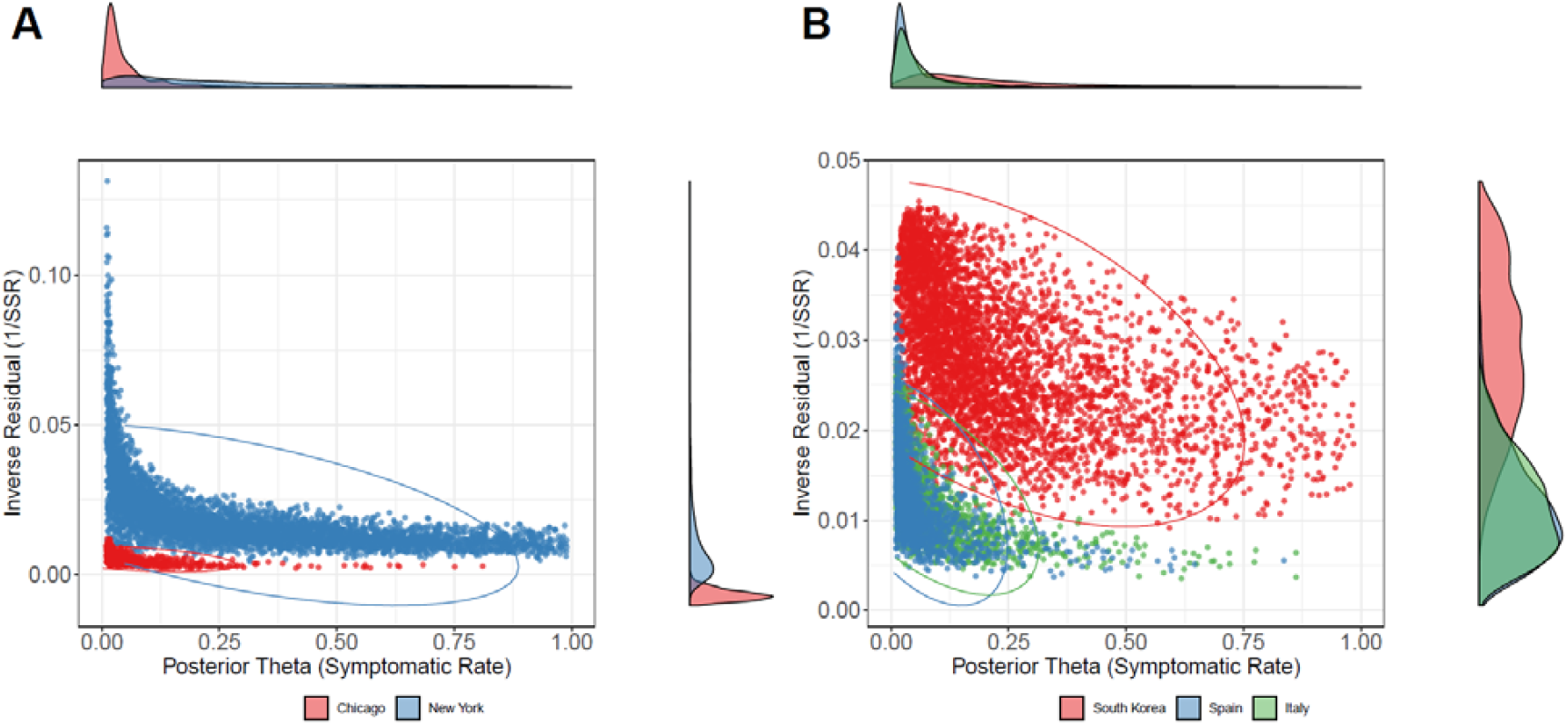
Scatterplot of inverse residual (goodness of fit) versus posterior sample of symptomatic rate. The posterior distribution of symptomatic rate *θ* and the inverse of sum of squared residuals (SSR^-1^) for Chicago and New York City (A) and the same set of priors for Italy, Spain, and South Korea (B). The marginal posterior density of *θ* and SSR^-1^ are displayed on the right and top of the scatter plot, respectively. The ellipses encircle 95% of simulation runs for each location.

**Figure 4.**
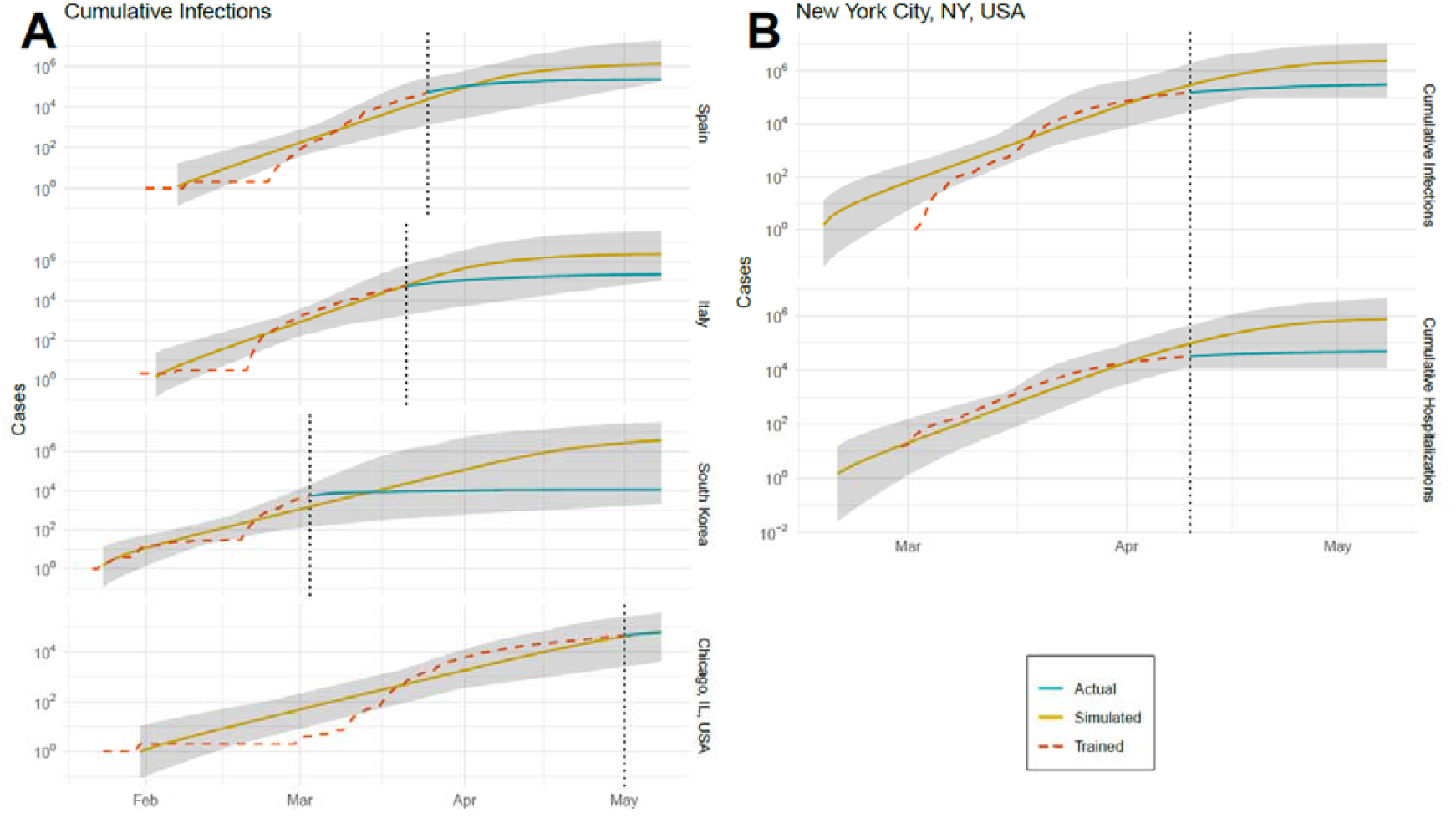
Simulated versus actual cumulative infections and hospitalization. (A) The simulated and actual cumulative infections for each location (Spain, Italy, South Korea, Chicago, and New York City are shown) for three different prior distributions of the symptomatic rate (*θ*). The turquoise line (simulated) corresponds with the simulated outputs for cumulative confirmed cases, which were fitted to the yellow line (trained) representing the actual data. The red dashed line represents the actual data after the explosion date, which is demarcated by the vertical black dotted line. The grey ribbon represents the upper and lower bounds of the MCMC sampling for all cases for each day.(B) Simulated and actual cumulative hospitalizations and cumulative infections for New York City.

## Discussion

With no end in sight to the ongoing COVID-19 pandemic, understanding the rate of disease spread and the extent of people infected is vital to designing effective containment policies. This analysis contributes to available disease models, which play an integral role in defining the policy choices of government officials, by providing accurate evidence that conforms to observed data. Given the growth rate in infection cases in the exponential phase and assuming the force of infection for undetected cases is lower, our results for all scenarios and countries suggest that the undetected population is significantly larger than the symptomatic population, the latter of which accounts for only 3-25% of the total infected population. The value of *R*_*0*_ varies from 2·8-3·6, which corresponds with a growing consensus that *R*_*0*_ is approximately 3·1 (Table 1). The variation in symptomatic cases is almost certainly a function of testing, which has been quite variable among cities and countries, yet even in South Korea, which had extremely high rates of testing during the beginning of the pandemic, our models still suggest large fractions of cases were likely missed due to asymptomatic/mild infections. Nevertheless, higher testing rates in South Korea also led to a slower transmission of SARS-CoV-2. This is also true when you compare testing in New York City versus Chicago, where testing per capita during the exponential phase was higher in New York City.

If the proportion of individuals that are asymptomatically infected is higher than initially assumed, policies targeting symptomatic individuals, such as travel restrictions on affected areas or quarantines of sick individuals, are not productive. By the time these policies can be implemented, a large proportion of the population may already be infected but not yet infectious due to the long incubation period [35]. Evidence from China suggests that the observed infection data lagged reported infections by about two weeks -- despite a dramatic drop in transmission between January 15 and January 25, the rate of newly confirmed cases did not begin to level off country-wide and within each local city until two weeks later. As of May 18, 2020, the uptick of COVID-19 cases has forced northeast China to reinitiate lockdown measures [36], which indicates the sensitivities and caution around case reemergence. Accurate assessments of the risk of community spread are needed to inform these decisions about when and where to re-impose restrictions to contain the spread of SARS-CoV-2. Our study found that asymptomatic cases may be easily missed without extensive testing, leading to the continued, undetected spread of disease despite possibly lower transmission rates among asymptomatic individuals compared to more severe cases. Thus, identifying asymptomatic cases is imperative for reducing widespread transmission and explosive growth of the disease, especially if a large majority of the population remains susceptible.

A potentially large asymptomatically infected population has important implications for policy regarding herd immunity. In recent reports, the vast majority of available evidence suggests that infected individuals develop some level of immunity to circulating strains of SARS-CoV-2, but there are potential differences in immunity between symptomatic and asymptomatic cases [37]. As more of the population gains immunity, the susceptible population will decrease and disease spread will slow. When transmissibility between contacts falls because of widespread immunity, the effective reproductive number is reduced, the disease spreads more slowly, and the threat posed by widespread numbers of infected individuals fades away. Such widespread immunity would allow restrictions to be lifted sooner, as most individuals would not pose a transmission risk to the general public and would abrogate the possibility of a second peak in the future. Our analysis underscores the likelihood of a large, hitherto undetected population of immune individuals in areas hit hard by the pandemic. Widespread, representative serological surveys are therefore essential to understanding the extent of disease spread and the necessity of mandatory social distancing policies to contain the spread of the virus. Understanding the potential risks requires serological surveys to be conducted as soon as possible across representative populations of disease burden to improve our understanding of disease transmission and to inform better policies regarding quarantines.

As with most COVID-19 modeling studies, the limitations of epidemiological models have been constrained by data and testing quality regarding the true prevalence of SARS-CoV-2 infections. We assumed for the beginning stages of the pandemic, the confirmed cases only reflected cases that were mostly moderate and severely symptomatic, while asymptomatic and mild cases were mostly overlooked due to the limited supply of testing kits. The uncertainty of these testing differences was implicitly captured in the Bayesian framework. The population size for each country are simplified and assumed to be static. Nevertheless, the parsimonious model provides a conservative estimate of undetected cases since traveling individuals make up a negligible proportion population, and if these travelers are infected and undetected in early stages of the pandemic, it will further support our claim. Furthermore, the estimation of transmission is assumed to be homogenous population mixing, while in reality contact networks are heterogeneous with varying contact patterns.

## Data Availability

Data derived from public domain resources

https://github.com/CSSEGISandData/COVID-19

## Supporting information

**S1 Appendix. Model Specifications, Deriving the Next Generation Matrix and Calculating R0, Bayesian Inference of Model Parameters, Population Inclusion for US Cities, and Supplement Plots**

