## Supplementary material for "Modeling the “Bomb-Like” Dynamics of COVID-19 with Undetected Transmissions and the Implications for Policy": S1 Appendix

### <u>S1 Appendix</u>. Model Specifications, Deriving the Next Generation Matrix and Calculating $R_0$, Bayesian Inference of Model Parameters, Population Inclusion for US Cities, and Supplement Plots

Supplement to:

#### Modeling the "Bomb-Like" Dynamics of COVID-19 with Undetected Transmissions and the Implications for Policy

Gary Lin, Anindya Bhaduri, Alexandra T. Strauss, Maxwell Pinz, Diego A. Martinez, Katie K. Tseng, Oliver Gatalo, Andrew T. Gaynor, Efrain Hernandez-Rivera, Emily Schueller, Yupeng Yang, Simon A. Levin, Eili Y. Klein, For the CDC MInD-Healthcare Program

#### 1. Model Specifications

Using the classic SIR model formulation from Kermack and McKendrik, we utilized the classical epidemiological dynamics to reflect viral transmission patterns of the COVID-19. We defined five compartmental states: susceptible (S), exposed (E), asymptomatic and mild symptomatic infections (C), moderate to severe symptomatic infections (I), and recovered (R), which are described in the main text. The model can be formulated as a set of ordinary differential equations (ODE).

$$
\begin{aligned}
&\dot{S} = -\alpha\beta\frac{SC}{N} - \beta\frac{SI}{N} \\
&\dot{E} = \alpha\beta\frac{SC}{N} + \beta\frac{SI}{N} - \mu E \\
&\dot{C} = \mu E(1-\theta) - \gamma_1 C \\
&\dot{I} = \mu\theta E - \gamma_2 I \\
&\dot{R} = \gamma_1 C + \gamma_2 I
\end{aligned} \tag{1}
$$

Transmission rate due to symptomatic population, $\beta$, is assumed to be reduced by α for asymptomatic transmissions. The detected symptomatic rate, $\theta$, determines the fraction of exposed individuals that transition to the infected state with moderate and severe symptoms. The inverse of the incubation period is defined by $\mu$, and the inverse of the infectiousness period for infected individuals *I* with moderate to severe symptoms group and infected group *C* with mild or no symptoms are indicated by $\gamma_1$ and $\gamma_2$, respectively.

The initial susceptible population is assumed to be the population size of the country based on population estimates from the World Bank [1]. The simulated infections with moderate to severe symptoms (*I*) was fitted to data on confirmed cases reported by the Center for System Science and Engineering at Johns Hopkins University [2]. For New York City, we fitted the model to cumulative hospitalizations collected by the New York State Department of Health [3], as well as cumulative confirmed cases. Therefore, it was necessary to divide the *I* compartment such that it is portioned into hospitalized, $I_H$, and symptomatic, $I_N$. This required us to include additional parameters, $\gamma_3$, recovery rate of hospitalizations, and $\varphi$, hospitalization rate. Hence, the augmented model with hospitalization is,

$$
\begin{aligned}
&\dot{S} = -\alpha\beta\frac{SC}{N} - \beta\frac{SI}{N} \\
&\dot{E} = \alpha\beta\frac{SC}{N} + \beta\frac{SI}{N} - \mu E \\
&\dot{C} = \mu E(1-\theta) - \gamma_1 C \\
&\dot{I_N} = \mu\theta E(1-\varphi) - \gamma_2 I_N \\
&\dot{I_H} = \varphi\mu\theta E - \gamma_3 I_H \\
&\dot{R} = \gamma_1 C + \gamma_2 I_N + \gamma_3 I_H
\end{aligned} \tag{2}
$$

In order to initialize the model, we assume the first-time step of the simulation is the same as the first confirmed case for each country, in which we seed only one infected person in the simulation. In the simulation, we added an additional parameter that is used for estimating. For Italy, Spain, South Korea, and Chicago we seeded the simulation with one infected person, respectively, on January 31st, 2020, February 1st, 2020, January 22nd, 2020, and January 22nd, 2020. For New York, we assumed there was a relatively longer delay in reporting, so we started the simulation 14 days before the first confirmed case on February 17, 2020. Since we were interested in the exponential growth of infection, the simulation time window ended when the country reached an inflection point where social distancing and lockdown effects were observed. Therefore, we ended the simulation on March 21st, 2020, March 25th, 2020, March 3rd, 2020, April 22nd, 2020, and April 10th, 2020 for Italy, Spain, South Korea, Chicago, and New York City. The set of ordinary differential equations (ODEs) were solved using the LSODA algorithm [4], and implemented in Python 3·8.

#### 2. Deriving the Next Generation Matrix and Calculating $R_0$

To assess the biological relevance of the parameters, we calculated the $R_0$ of the model, based on the dominant eigenvalue of the *next generation matrix*. The infection subsystem of the model can be described by equations (2).

(3)

$$\dot{E} = \alpha\beta\frac{SC}{N} + \beta\frac{SI}{N} - \mu E$$

$$\dot{C} = \mu E(1-\theta) - \gamma_1 C$$

$$\dot{I} = \theta\mu E - \gamma_2 I$$

We use a linearization of the infection subsystem, similar to the formulation outlined by Diekmann et al. [5],

(4)

$$\dot{\mathbf{x}} = (\mathbf{T} + \mathbf{M})\mathbf{x}$$

where **x** = [E, C, I], $T$ represents the transmission matrix that captures the number of transmissions from the susceptible compartments, and the matrix M represents the transition between compartments within the infection subsystem. Specifically, $T$ is equal to

$$\mathbf{T} = \begin{bmatrix} 0 & \alpha\beta & \beta \\ 0 & 0 & 0 \\ 0 & 0 & 0 \end{bmatrix}$$

and M is equal to

$$\mathbf{M} = \begin{bmatrix} -\mu & 0 & 0 \\ (1-\theta)\mu & -\gamma_1 & 0 \\ \theta\mu & 0 & -\gamma_2 \end{bmatrix}.$$

From this formulation, we can mathematically construct the next generation matrix $W$**,** which is defined as

$$\mathbf{W} = -\mathbf{TM}^{-1},$$

which would equate to

$$\mathbf{W} = -\begin{bmatrix} 0 & \alpha\beta & \beta \\ 0 & 0 & 0 \\ 0 & 0 & 0 \end{bmatrix} \cdot \begin{bmatrix} -\frac{1}{\mu} & 0 & 0 \\ -\frac{(1-\theta)}{\gamma_1} & -\frac{1}{\gamma_1} & 0 \\ -\frac{\theta}{\gamma_2} & 0 & -\frac{1}{\gamma_2} \end{bmatrix}$$

$$= \begin{bmatrix} \alpha\beta\frac{(1-\theta)}{\gamma_1} + \beta\frac{\theta}{\gamma_2} & \alpha\beta\frac{1}{\gamma_1} & \beta\frac{1}{\gamma_2} \\ 0 & 0 & 0 \\ 0 & 0 & 0 \end{bmatrix}.$$

*R*$_0$ is equal to the dominant eigenvalue of **W** (highlighted in red), which is equal to

$$R_0 = \alpha\beta\frac{(1-\theta)}{\gamma_1} + \beta\frac{\theta}{\gamma_2}.$$

#### 3. Bayesian Inference of Model Parameters

In order to understand the credibility of parameters, Θ, we used Monte Carlo Markov Chain (MCMC) methods to estimate the P($\Theta \,|\, \hat{X}$), the posterior probability of the parameters, in which we are estimating based on the observed confirmed cases, $\hat{X}$. From the observed data, we were able to estimate the posterior distribution using the following Bayesian framework,

$$P(\Theta|\hat{X}) = \frac{P(\hat{X}|\Theta)P(\Theta)}{P(\hat{X})} \propto \mathcal{L}(\Theta)P(\Theta)$$

where $\mathcal{L}(\Theta) = P(\hat{X}|\Theta)$ is the likelihood function and P(Θ) is prior on our belief of Θ. The prior assumptions are shown in Tables 2 and 3 in the main text. Since we are assuming that the error follows a normal distribution $\mathcal{N}(0, \sigma^2)$ with known variance $\sigma^2$, the loglikelihood can be defined as

$$\log \mathcal{L}(\Theta) = -\log(2\pi\sigma^2) - \frac{SSR(\Theta)}{2\sigma^2}$$

where SSR is the sum of squared residuals between the simulated data $X_{it}$ and observed data $\hat{X}_{it}$. For Italy, Spain, South Korea, and Chicago, we fitted the model to confirmed cases. For New York City, we fitted the model to confirmed cases and cumulative hospitalizations, which means there are two times series that the MCMC is using to determine the goodness of fit. The goodness of fit is based on the sum of squared residuals (SSR) which is commonly used in time series analysis and can be calculated as

$$SSR = \sum_{i \in M} \sum_{t=0}^{T} \left(\log(X_{it}(\Theta) + 1) - \log(\hat{X}_{it} + 1)\right)^2$$

where the subscripts $i \in M$ represents a fitted series (e.g. cumulative infections), and $t$ represents a time point. The inverse sum of squared residuals, $SSR^{-1}$, are shown in Tables S1 and S2. We used the log value of the simulated and real data values to better capture the exponential increase. The infection evolution based on the fit are shown in Figures S20 and S21.

Using the Metropolis-Hasting (M-H) Algorithm, the posterior marginal distributions for each parameter in Θ were generated assuming uniform priors. For the uniform prior of the maximum rate of asymptomatic transmission parameter θ, three different set of bounds were considered: $0{\cdot}01 < \theta < 0{\cdot}10$, $0{\cdot}01 < \theta < 0{\cdot}50$, and $0{\cdot}01 < \theta < 0{\cdot}99$. Normally distributed proposal densities were considered to generate candidate samples for the acceptance-rejection step of the M-H algorithm. Each MCMC simulation was run 400,000 times with parameter sampling originating from these priors. In order to obtain independent samples of the posterior distribution, a burn-in period of 10,000 samples, and thinning was conducted with appropriate intervals for each case. The ACF plots for all accepted parameters after thinning are shown in Figures S1-S15, and the trace plots are located in Figures S16-S30. The simulations were performed on the US Army Research Laboratory's Centennial HPC system across 160 compute cores. These cores are rated at 2·2 GHz and 128 GBs.

From the posterior distribution, we were able to calculate the 95% credible interval (95% CrI) of each posterior distribution of the parameters based on the median values. Tables S1 and S2 show a complete listing of the posterior estimation for all parameters.

#### 4. Population Inclusion for US Cities

For Chicago and New York City we included the population defined by the Metropolitan statistical area (MSA), which included the following counties and their associated Federal Information Processing Standards code (FIPS):

**Chicago, IL**

| ***County*** | ***FIPS*** |
| --- | --- |
| Cook County | 17031 |
| DuPage County | 17043 |
| Grundy County | 17063 |
| Kendall County | 17093 |
| McHenry County | 17111 |
| Will County | 17197 |

**New York City, NY**

| ***County*** | ***FIPS*** |
| --- | --- |
| Bergen County | 34003 |
| Hudson County | 34017 |
| Middlesex County | 34023 |
| Monmouth County | 34025 |
| Ocean County | 34029 |
| Passaic County | 34031 |
| Orange County | 36071 |
| Rockland County | 36087 |
| Westchester County | 36119 |
| New York City | 36061 |

**Table S1.** Posterior parameters estimation for countries from the Bayesian analysis using the epidemiological model where the means and corresponding credible intervals are shown in the parenthesis.

| | | $\bar{\theta}$ < 10% | | | $\bar{\theta}$ < 50% | | | $\bar{\theta}$ < 99% (Unconstrained Prior) | | |
|---|---|---|---|---|---|---|---|---|---|---|
| Variable | Prior Distribution | Italy | Spain | South Korea | Italy | Spain | South Korea | Italy | Spain | South Korea |
| $R_0$ | -- | **3.05 (1.06-26.75)** | **3.34 (1.08-30.97)** | **2.02 (0.94-16.84)** | **3.43 (1.1-29.34)** | **3.54 (1.08-31.91)** | **2.49 (1-18.46)** | **3.25 (1.09-29.77)** | **3.62 (1.13-34.89)** | **2.75 (1.04-22.44)** |
| $\theta$ | **U(0.01, $\bar{\theta}$)**† | **0.03 (0.01-0.09)** | **0.03 (0.01-0.09)** | **0.06 (0.01-0.1)** | **0.05 (0.01-0.32)** | **0.03 (0.01-0.25)** | **0.14 (0.02-0.44)** | **0.04 (0.01-0.41)** | **0.03 (0.01-0.32)** | **0.18 (0.02-0.85)** |
| $\alpha$ | U(0.01,0.99) | 0.68 (0.26-0.97) | 0.68 (0.25-0.97) | 0.64 (0.21-0.97) | 0.67 (0.23-0.97) | 0.68 (0.23-0.97) | 0.6 (0.12-0.96) | 0.67 (0.24-0.97) | 0.68 (0.22-0.97) | 0.58 (0.09-0.96) |
| $\beta$ | U(0.01,2) | 1.34 (0.54-1.96) | 1.33 (0.52-1.96) | 1.32 (0.49-1.95) | 1.36 (0.55-1.96) | 1.36 (0.51-1.96) | 1.28 (0.45-1.95) | 1.38 (0.53-1.96) | 1.36 (0.51-1.96) | 1.29 (0.43-1.95) |
| $\gamma_1$ | U(0.01,0.99) | 0.21 (0.02-0.82) | 0.18 (0.02-0.78) | 0.34 (0.03-0.92) | 0.2 (0.02-0.82) | 0.18 (0.02-0.79) | 0.34 (0.03-0.93) | 0.22 (0.02-0.84) | 0.18 (0.02-0.78) | 0.33 (0.03-0.94) |
| $\gamma_2$ | U(0.01,0.30) | 0.49 (0.04-0.96) | 0.5 (0.04-0.96) | 0.47 (0.03-0.95) | 0.47 (0.03-0.95) | 0.49 (0.03-0.96) | 0.4 (0.03-0.95) | 0.49 (0.03-0.96) | 0.5 (0.03-0.96) | 0.37 (0.03-0.95) |
| $\mu$ | U(0.01,0.99) | 0.37 (0.1-0.93) | 0.29 (0.07-0.91) | 0.51 (0.13-0.96) | 0.31 (0.07-0.92) | 0.26 (0.06-0.91) | 0.31 (0.07-0.93) | 0.31 (0.07-0.92) | 0.26 (0.05-0.89) | 0.25 (0.05-0.92) |
| Inverse Sum of Squared Residuals ($SSR^{-1}$) | -- | 0.015 (0.008-0.025) | 0.014 (0.007-0.028) | 0.035 (0.019-0.044) | 0.013 (0.006-0.024) | 0.013 (0.006-0.028) | 0.03 (0.015-0.043) | 0.013 (0.006-0.024) | 0.013 (0.006-0.028) | 0.029 (0.014-0.042) |

**Table S2.** Parameters estimation for US cities from the Bayesian analysis using the epidemiological model where the means and corresponding credible intervals are shown in the parenthesis.

| Variable | Prior Distribution | $\bar{\theta}$ < 10% | | $\bar{\theta}$ < 50% | | $\bar{\theta}$ < 99% (Unconstrained Prior) | |
|---|---|---|---|---|---|---|---|
| | | New York City | Chicago | New York City | Chicago | New York City | Chicago |
| $\boldsymbol{R_0}$ | -- | **3.09 (1.73-19.71)** | **3.16 (1-32.75)** | **3.13 (1.69-18.16)** | **3.4 (1-33.41)** | **3.31 (1.69-20.55)** | **3.46 (1.01-34.41)** |
| $\boldsymbol{\theta}$ | **U(0.1, $\bar{\theta}$)**† | **0.04 (0.01-0.1)** | **0.03 (0.01-0.09)** | **0.15 (0.01-0.47)** | **0.03 (0.01-0.26)** | **0.25 (0.01-0.89)** | **0.03 (0.01-0.36)** |
| $\alpha$ | U(0.01,0.99) | 0.72 (0.3-0.97) | 0.64 (0.16-0.97) | 0.64 (0.14-0.97) | 0.64 (0.17-0.97) | 0.61 (0.09-0.97) | 0.61 (0.16-0.97) |
| $\beta$ | U(0.01,2) | 1.47 (0.68-1.97) | 1.25 (0.34-1.96) | 1.41 (0.61-1.95) | 1.25 (0.33-1.93) | 1.35 (0.58-1.96) | 1.27 (0.4-1.95) |
| $\gamma_1$ | U(0.01,0.99) | 0.31 (0.03-0.89) | 0.18 (0.02-0.81) | 0.35 (0.03-0.92) | 0.18 (0.02-0.8) | 0.36 (0.03-0.93) | 0.17 (0.02-0.85) |
| $\gamma_2$ | U(0.01,0.30) | 0.48 (0.04-0.96) | 0.5 (0.03-0.96) | 0.44 (0.03-0.95) | 0.51 (0.03-0.97) | 0.38 (0.03-0.95) | 0.5 (0.03-0.97) |
| $\gamma_3$ | U(0.01,0.30) | 0.49 (0.04-0.96) | -- | 0.47 (0.03-0.96) | -- | 0.46 (0.03-0.95) | -- |
| $\varphi$ | U(0.01,0.50) | 0.35 (0.13-0.49) | -- | 0.34 (0.13-0.49) | -- | 0.34 (0.13-0.49) | -- |
| $\mu$ | U(0.05,0.99) | 0.58 (0.18-0.96) | 0.15 (0.03-0.84) | 0.49 (0.02-0.83) | 0.13 (0.12-0.14) | 0.42 (0.1-0.94) | 0.12 (0.02-0.83) |
| Inverse Sum Squared Residuals ($SSR^{-1}$) | -- | 0.018 (0.009-0.032) | 0.006 (0.003-0.01) | 0.013 (0.008-0.027) | 0.005 (0.003-0.01) | 0.012 (0.007-0.026) | 0.005 (0.003-0.01) |

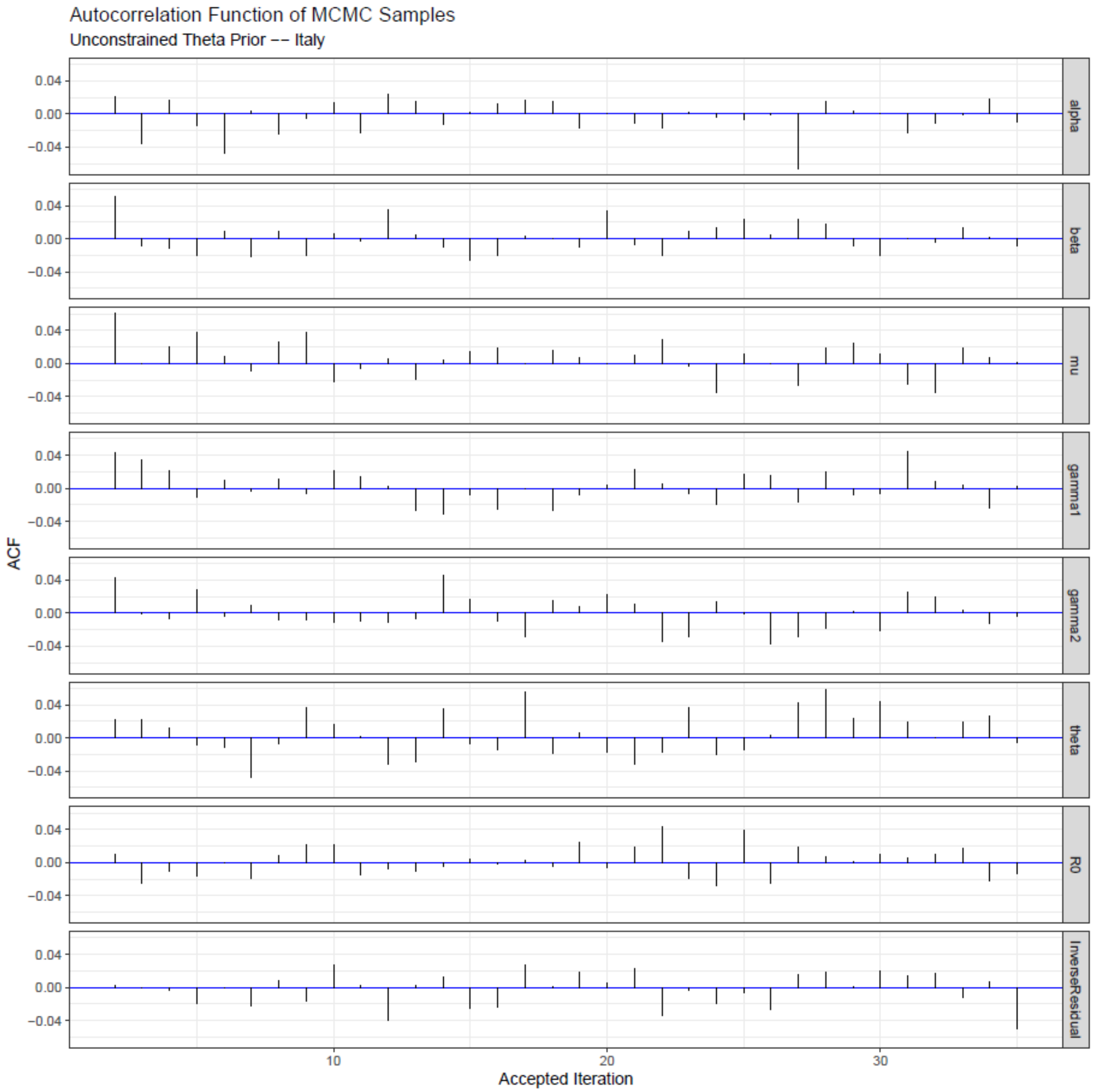

**Figure S1.** The autocorrelation of accepted parameters using the Metropolis-Hasting algorithm after thinning and burn-in are shown for Italy with unconstrained priors of θ.

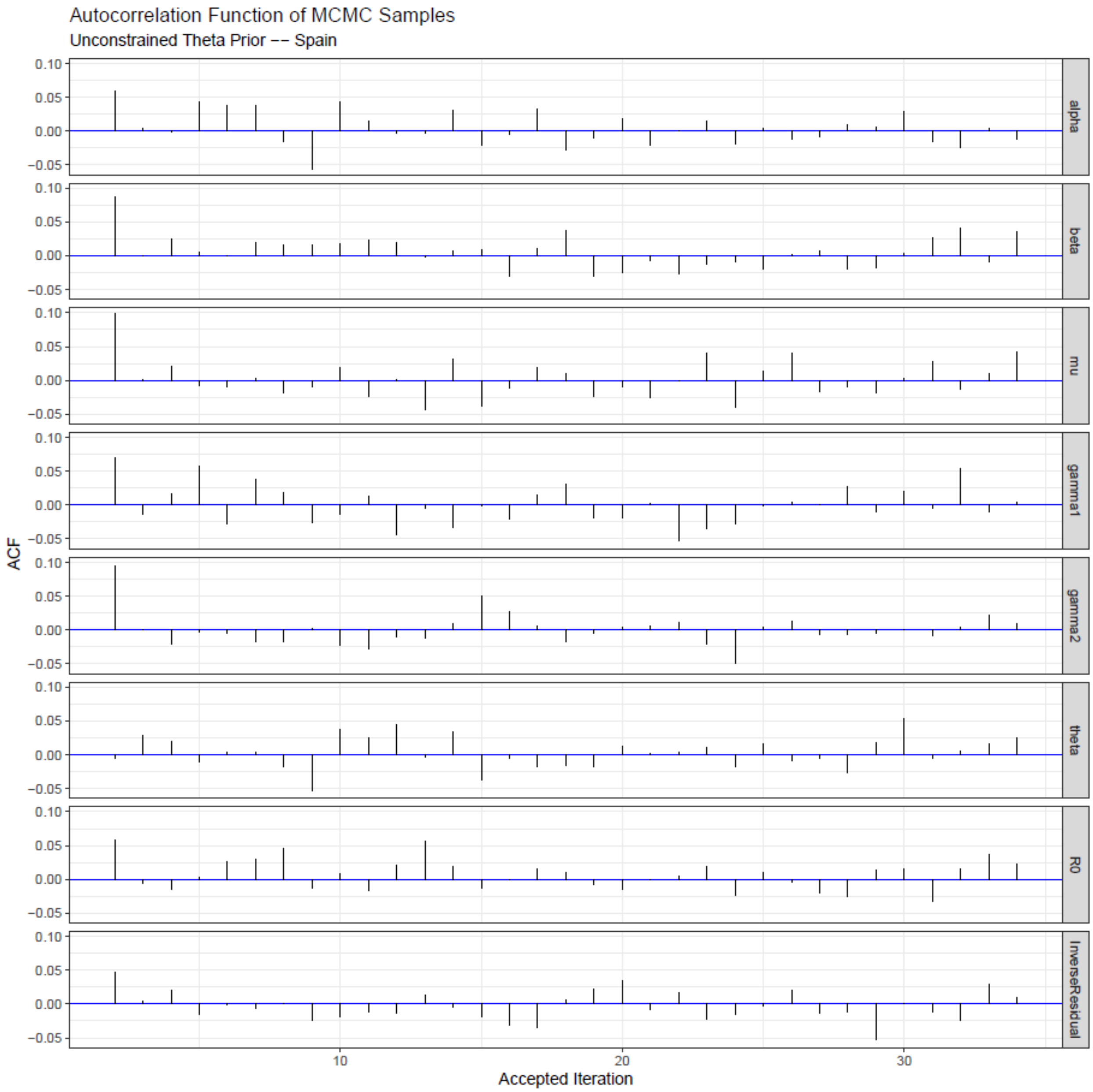

**Figure S2.** The autocorrelation of accepted parameters using the Metropolis-Hasting algorithm after thinning and burn-in are shown for Spain with unconstrained priors of θ.

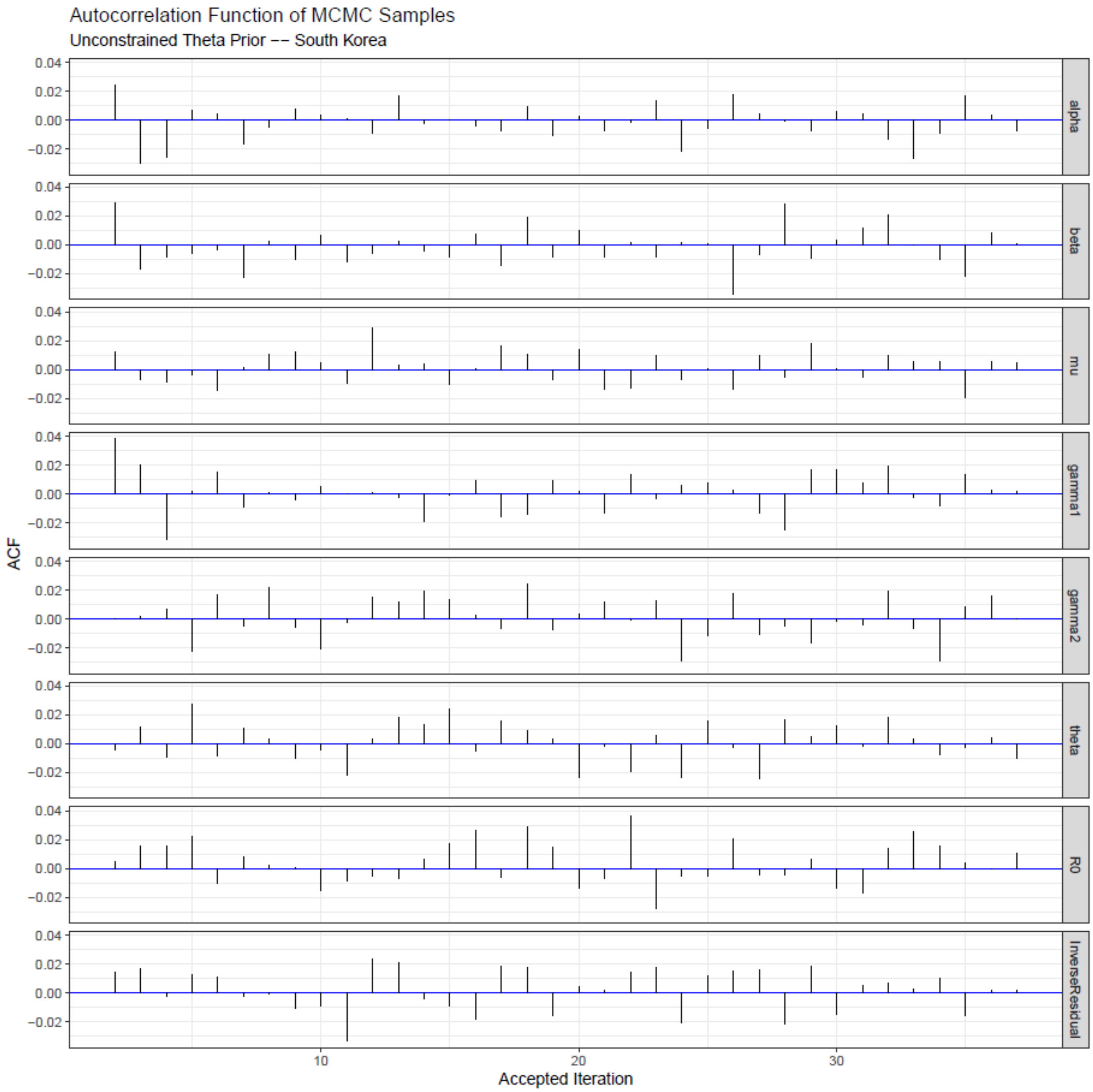

**Figure S3.** The autocorrelation of accepted parameters using the Metropolis-Hasting algorithm after thinning and burn-in are shown for South Korea with unconstrained priors of θ.

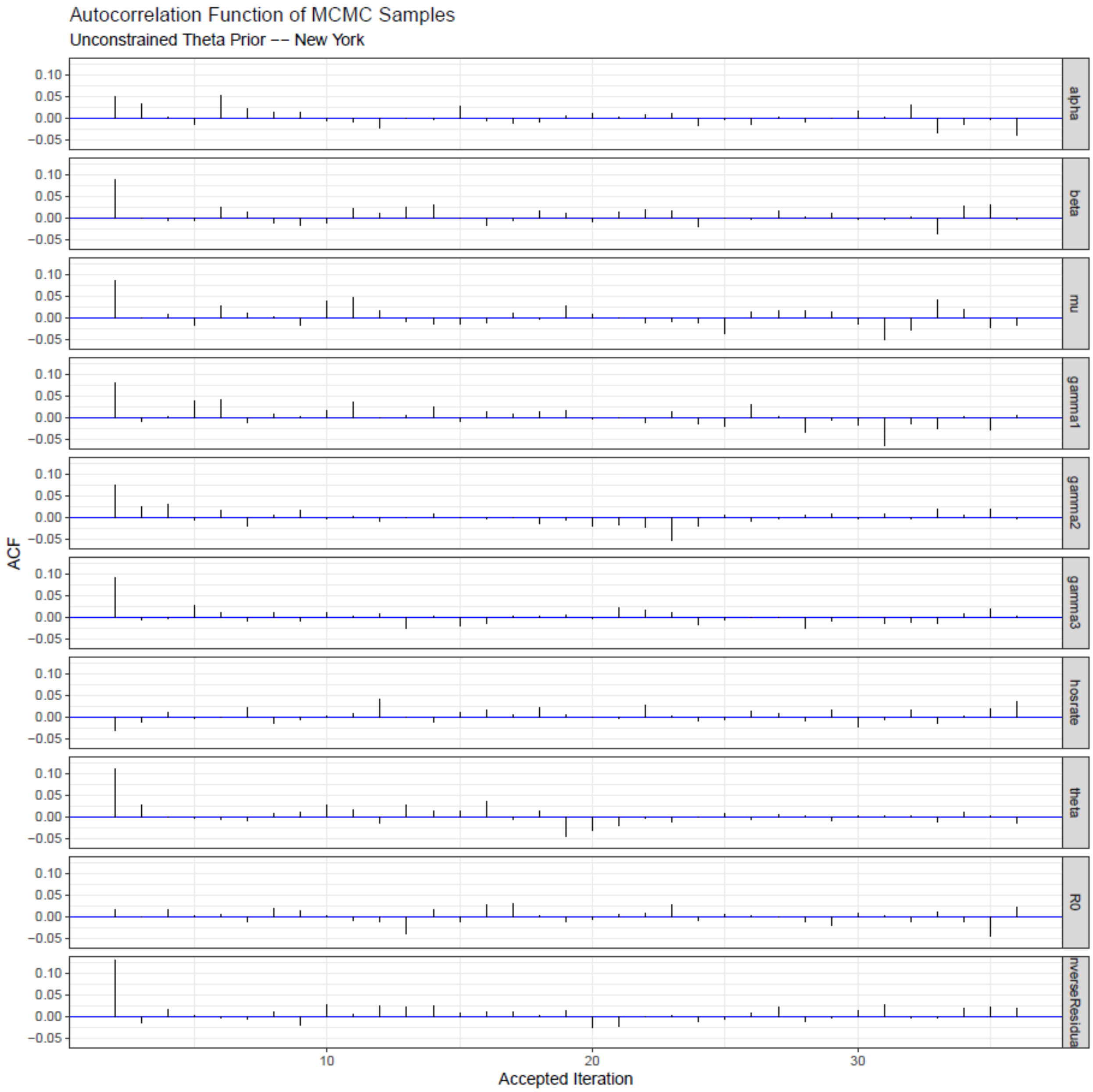

**Figure S4.** The autocorrelation of accepted parameters using the Metropolis-Hasting algorithm after thinning and burn-in are shown for New York City with unconstrained priors of θ.

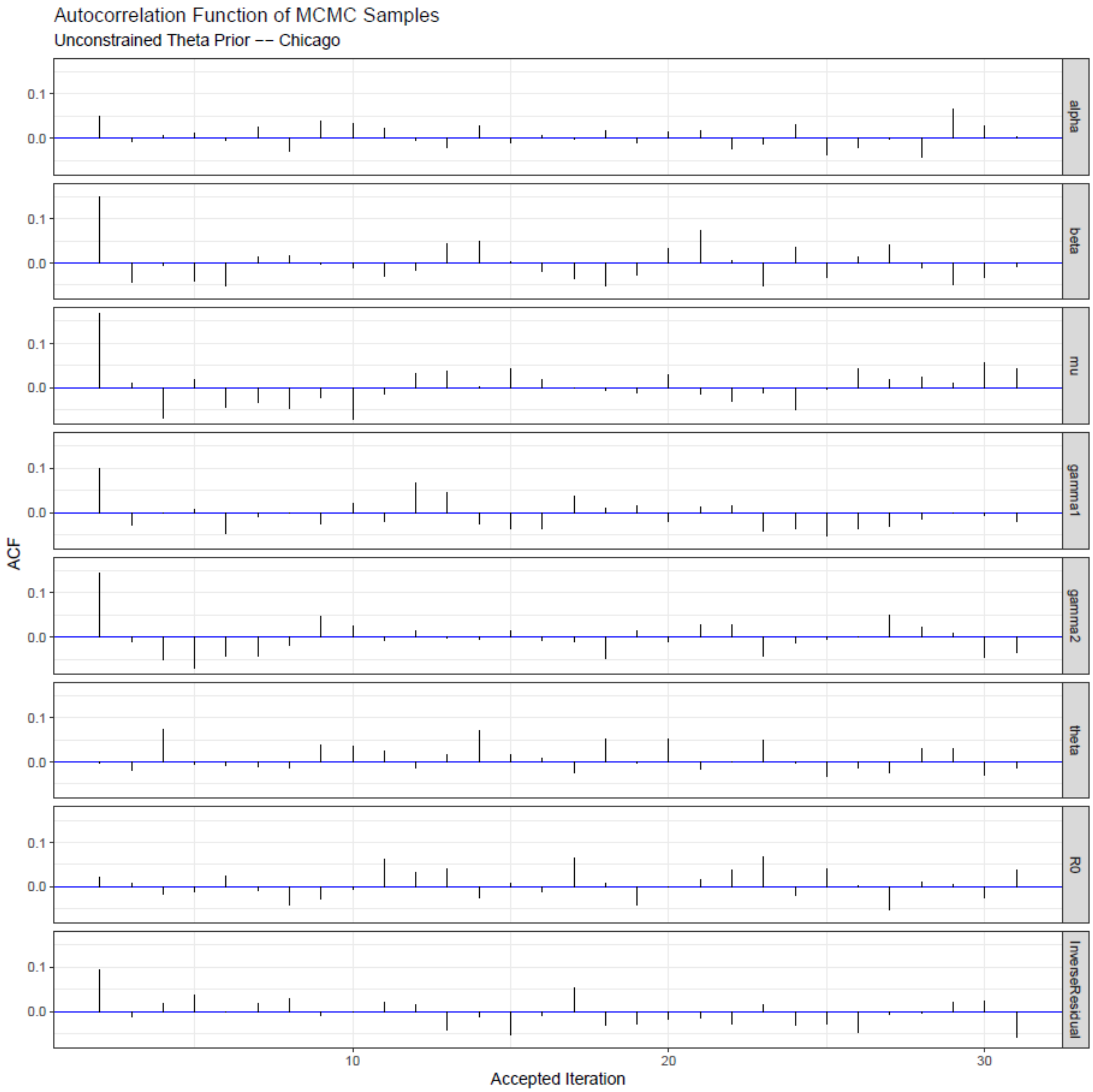

**Figure S5.** The autocorrelation of accepted parameters using the Metropolis-Hasting algorithm after thinning and burn-in are shown for Chicago with unconstrained priors of θ.

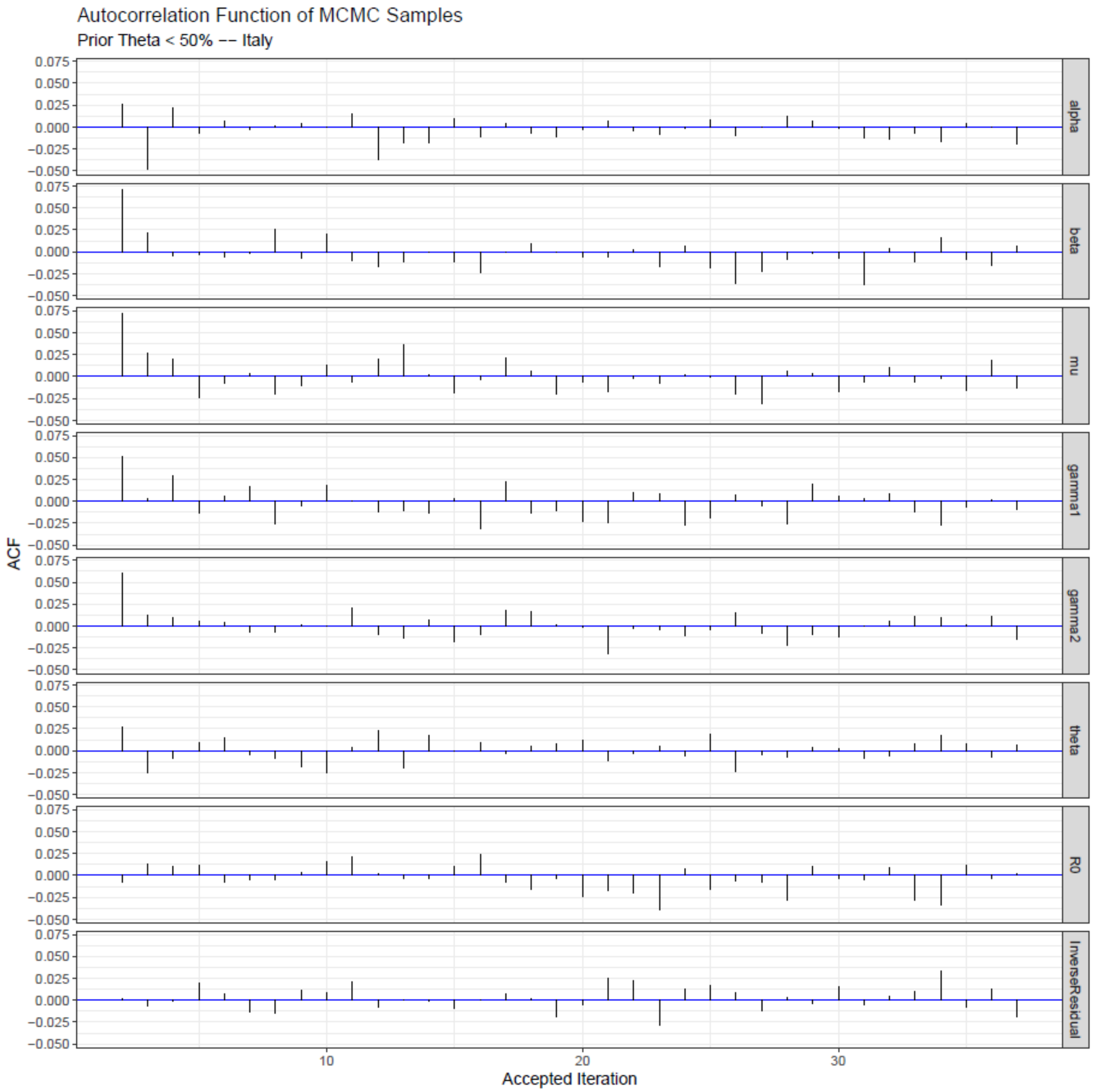

**Figure S6.** The autocorrelation of accepted parameters using the Metropolis-Hasting algorithm after thinning and burn-in are shown for Italy with priors of θ < 50%.

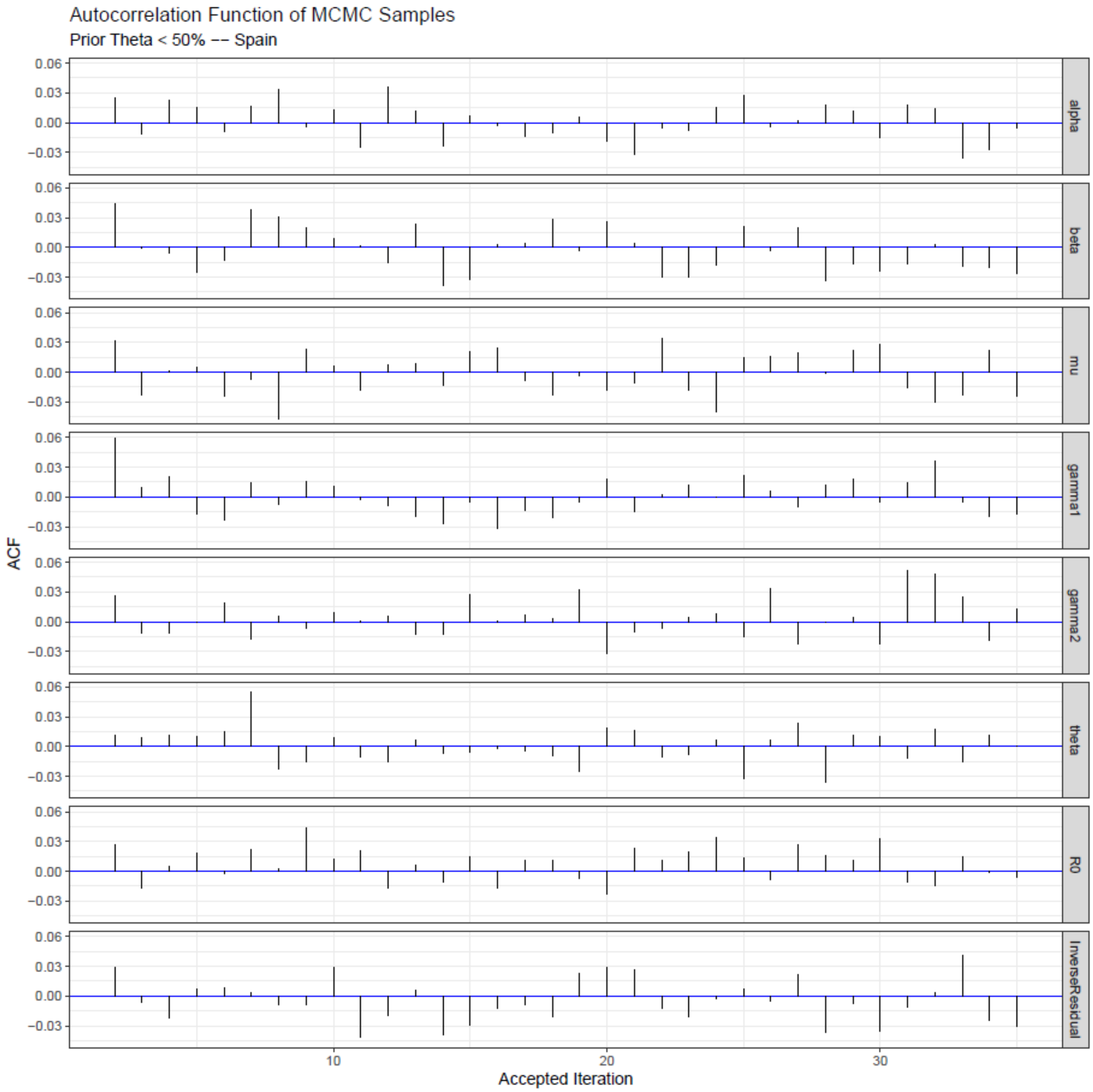

**Figure S7.** The autocorrelation of accepted parameters using the Metropolis-Hasting algorithm after thinning and burn-in are shown for Spain with priors of θ < 50%.

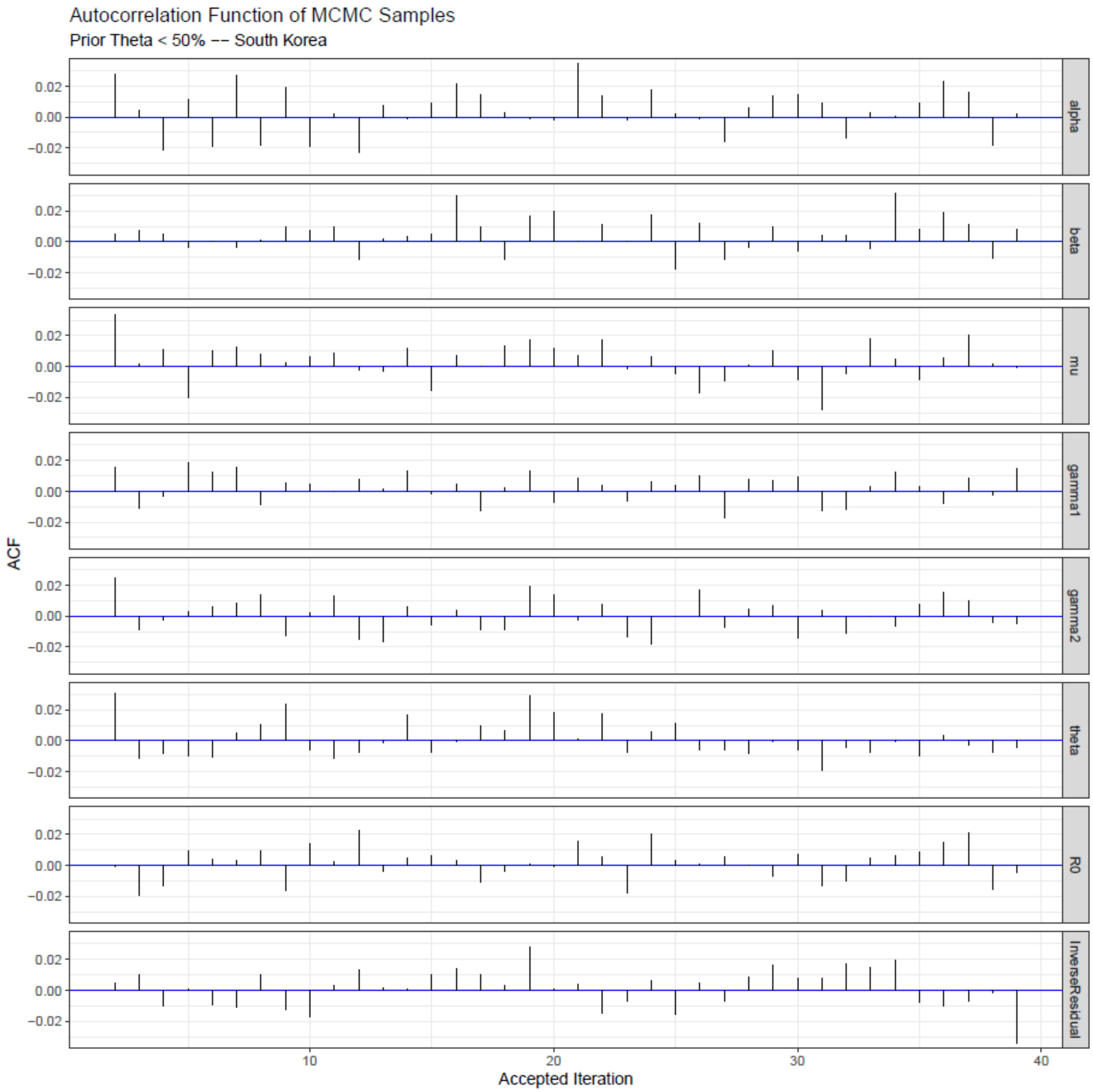

**Figure S8.** The autocorrelation of accepted parameters using the Metropolis-Hasting algorithm after thinning and burn-in are shown for South Korea with priors of $\theta < 50\%$.

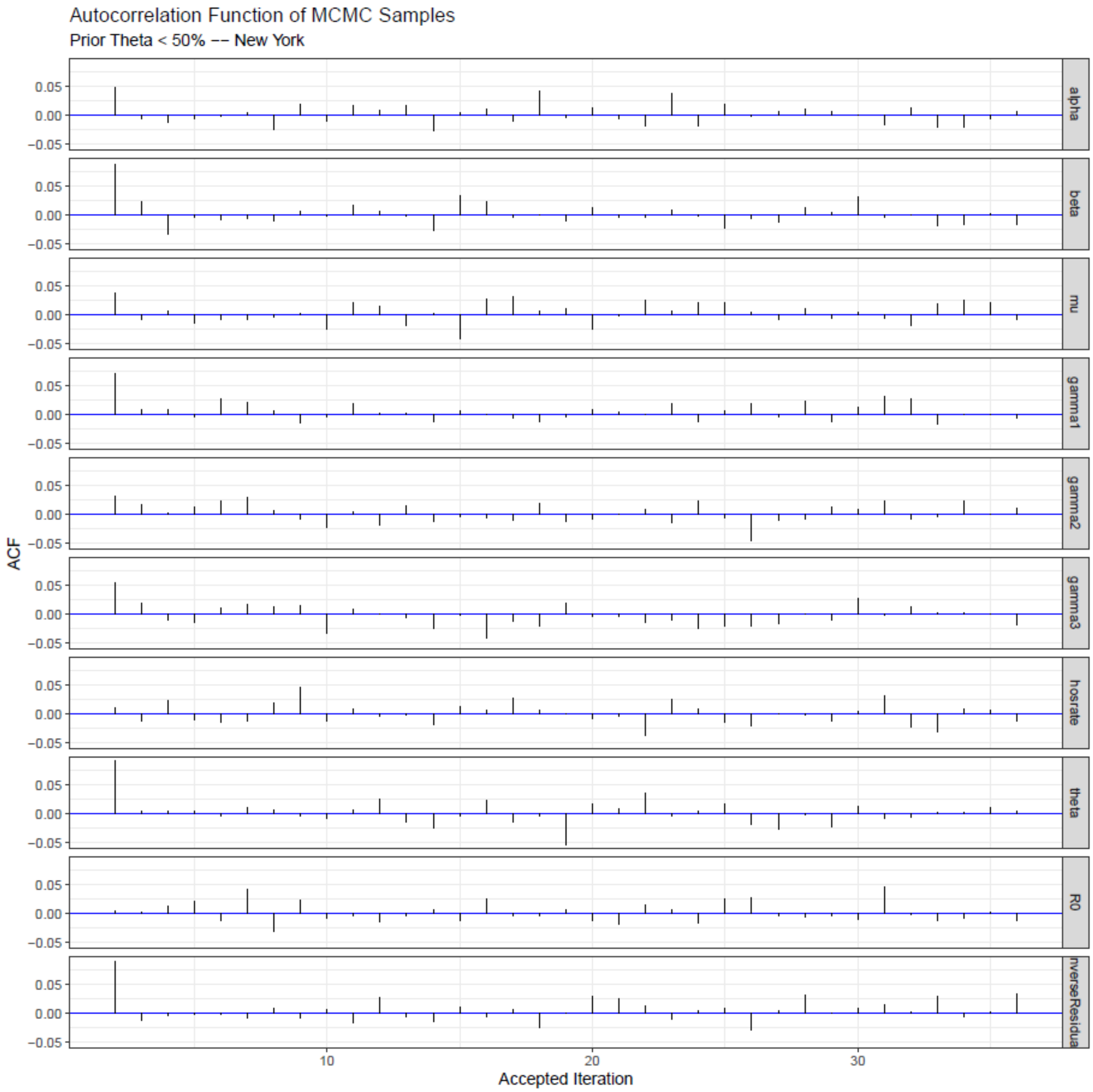

**Figure S9.** The autocorrelation of accepted parameters using the Metropolis-Hasting algorithm after thinning and burn-in are shown for New York City with priors of θ < 50%.

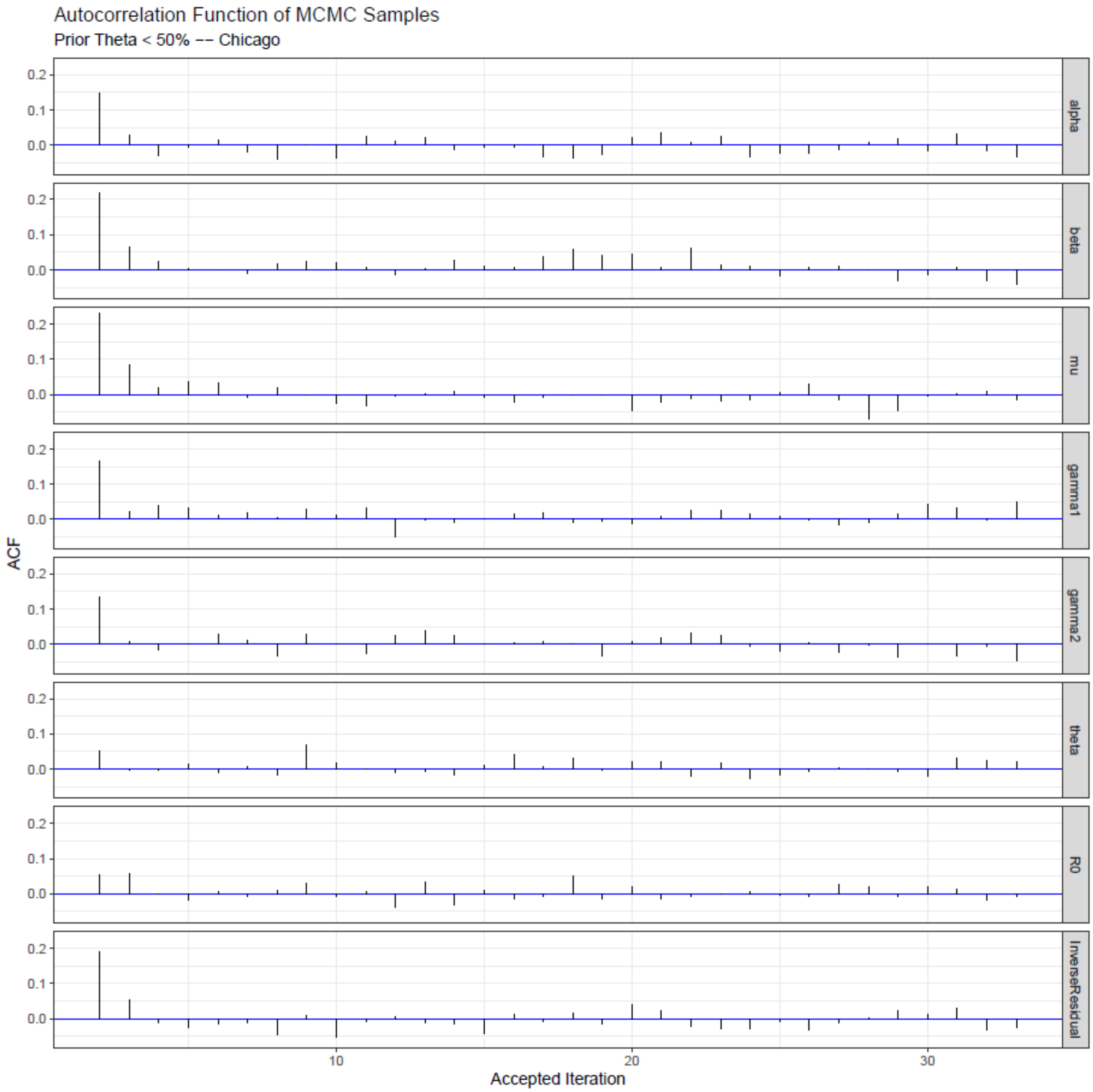

**Figure S10.** The autocorrelation of accepted parameters using the Metropolis-Hasting algorithm after thinning and burn-in are shown for Chicago with priors of θ < 50%.

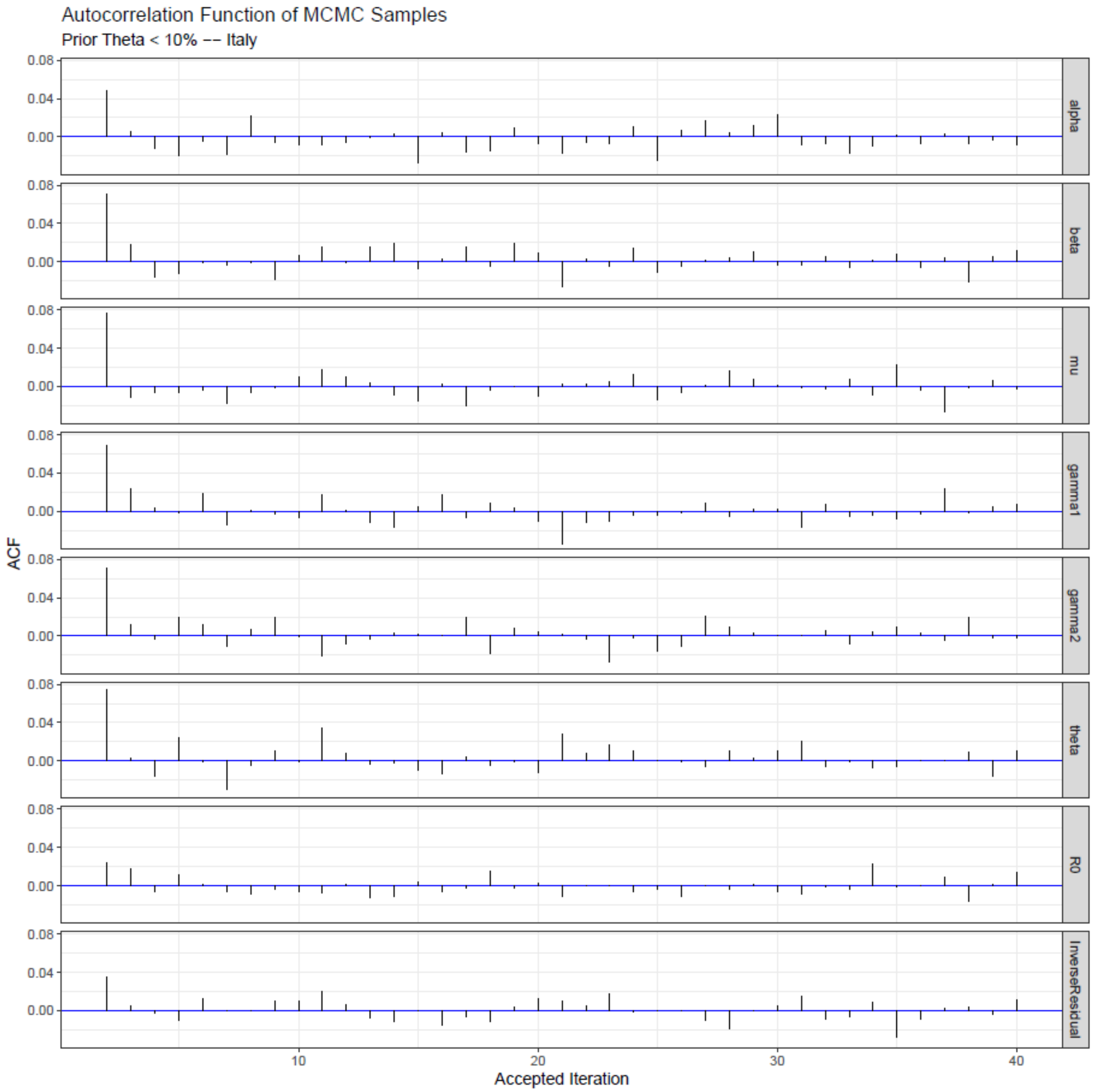

**Figure S11.** The autocorrelation of accepted parameters using the Metropolis-Hasting algorithm after thinning and burn-in are shown for Italy with priors of θ < 10%.

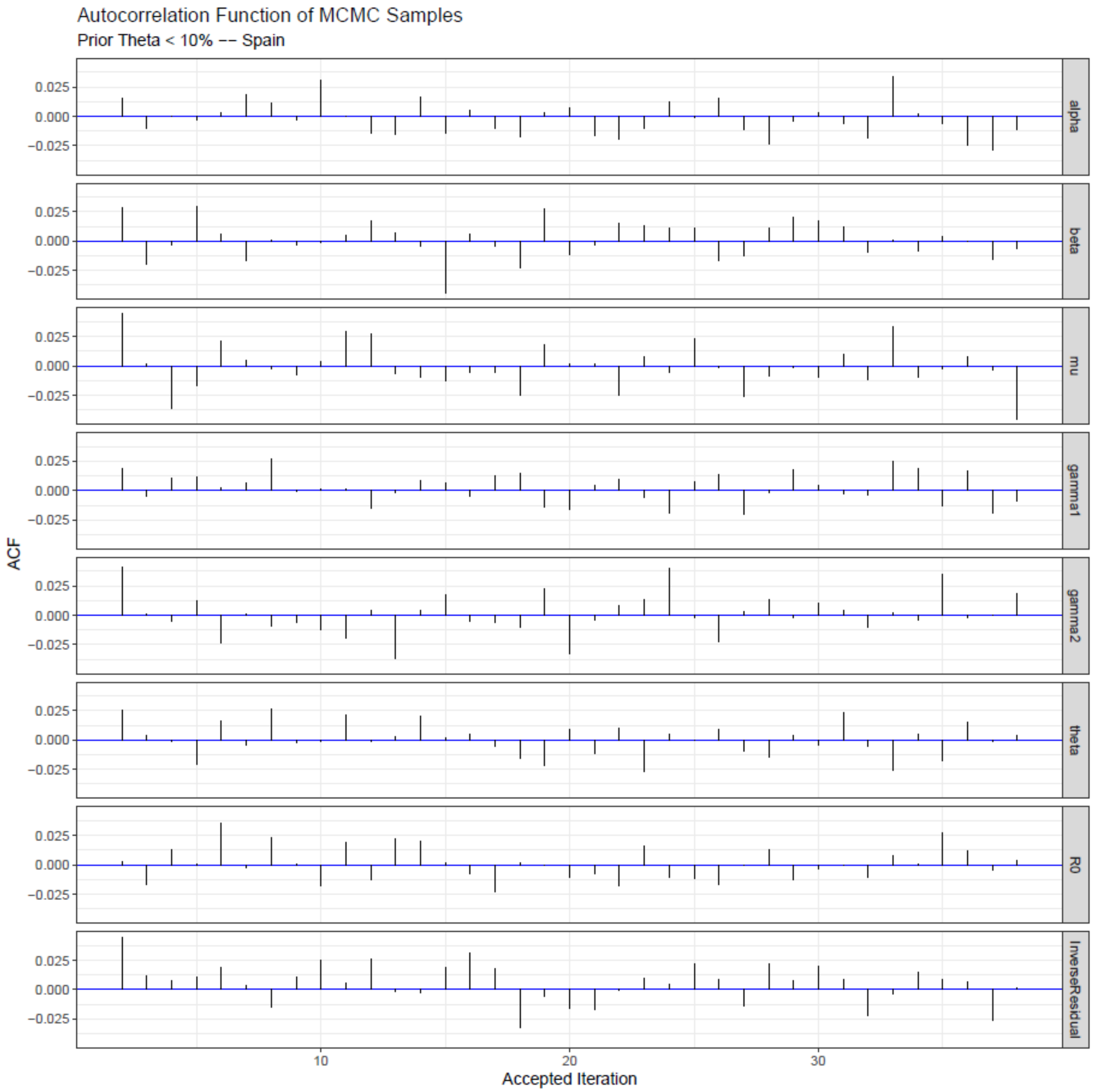

**Figure S12.** The autocorrelation of accepted parameters using the Metropolis-Hasting algorithm after thinning and burn-in are shown for Spain with priors of θ < 10%.

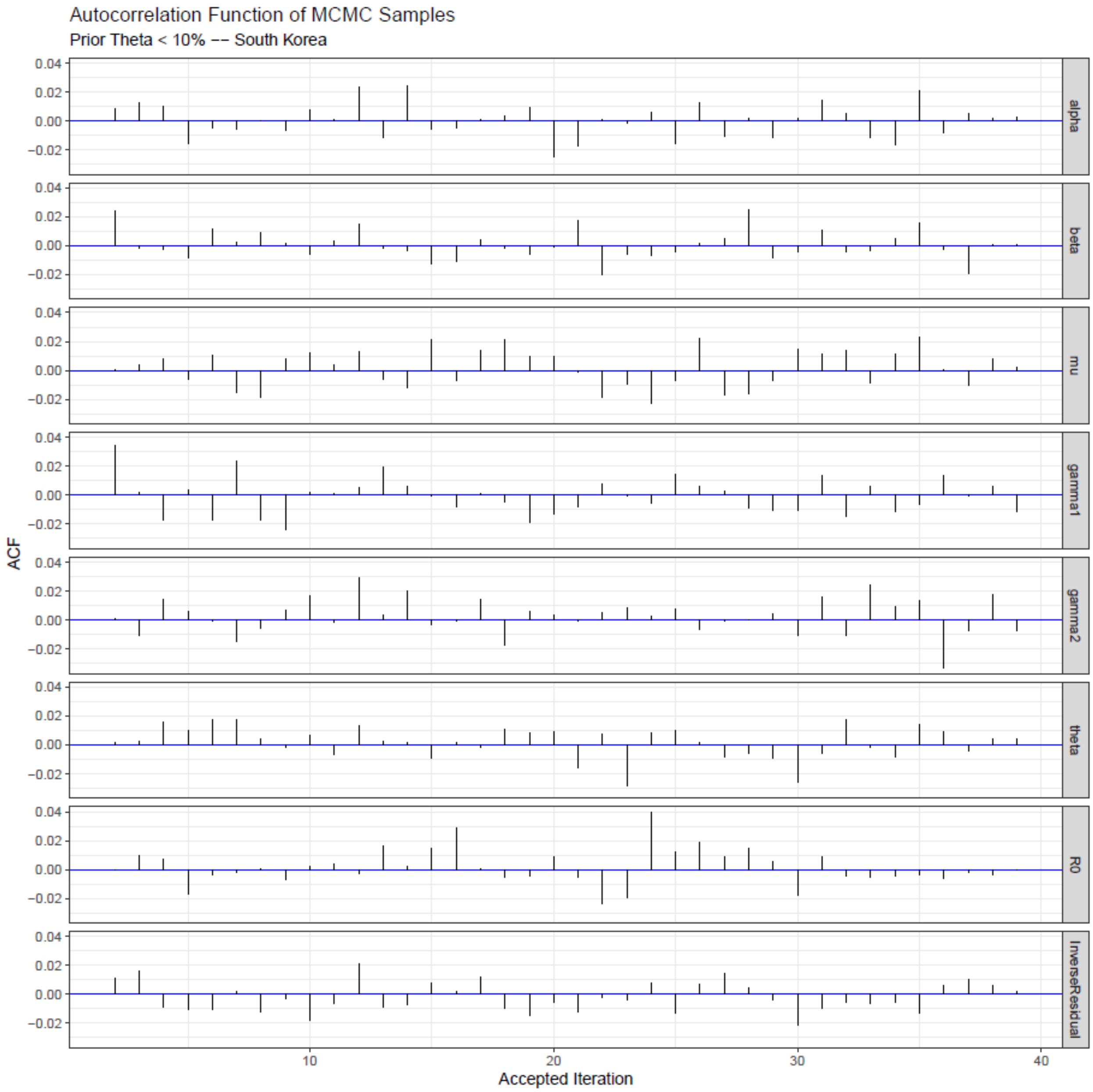

**Figure S13.** The autocorrelation of accepted parameters using the Metropolis-Hasting algorithm after thinning and burn-in are shown for South Korea with priors of θ < 10%.

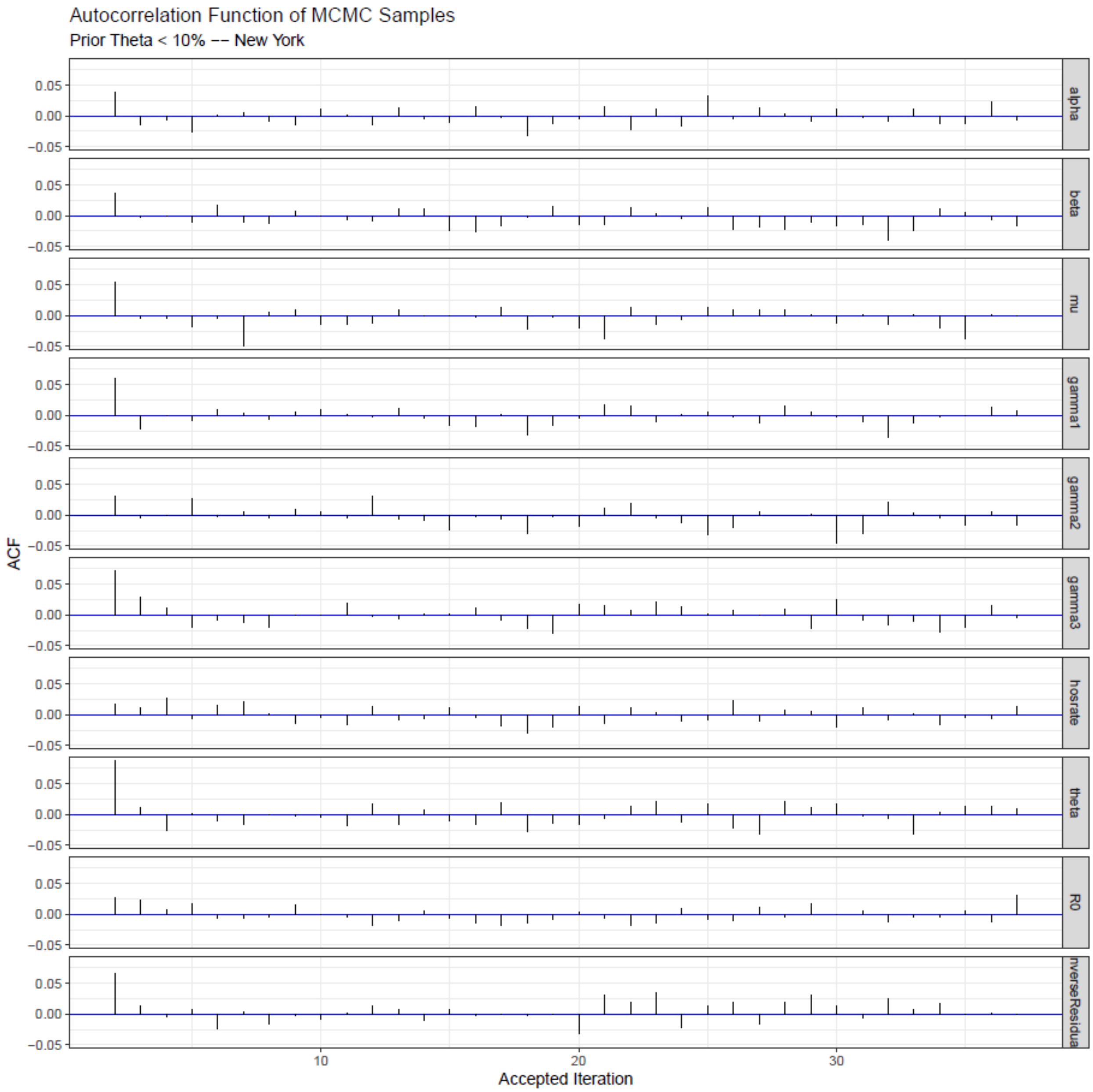

**Figure S14.** The autocorrelation of accepted parameters using the Metropolis-Hasting algorithm after thinning and burn-in are shown for New York City with priors of θ < 10%.

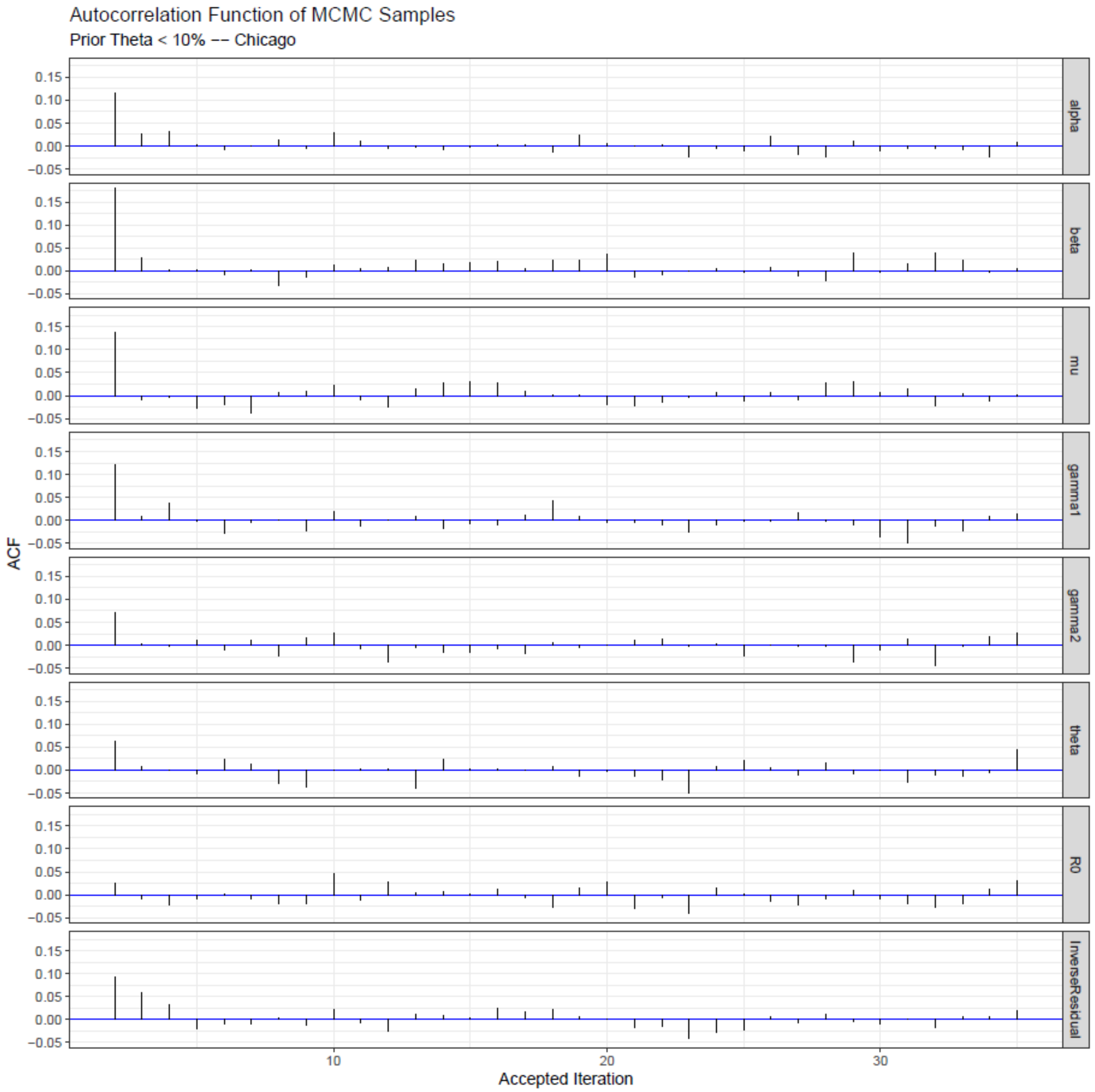

**Figure S15.** The autocorrelation of accepted parameters using the Metropolis-Hasting algorithm after thinning and burn-in are shown for Chicago with priors of $\theta < 10\%$.

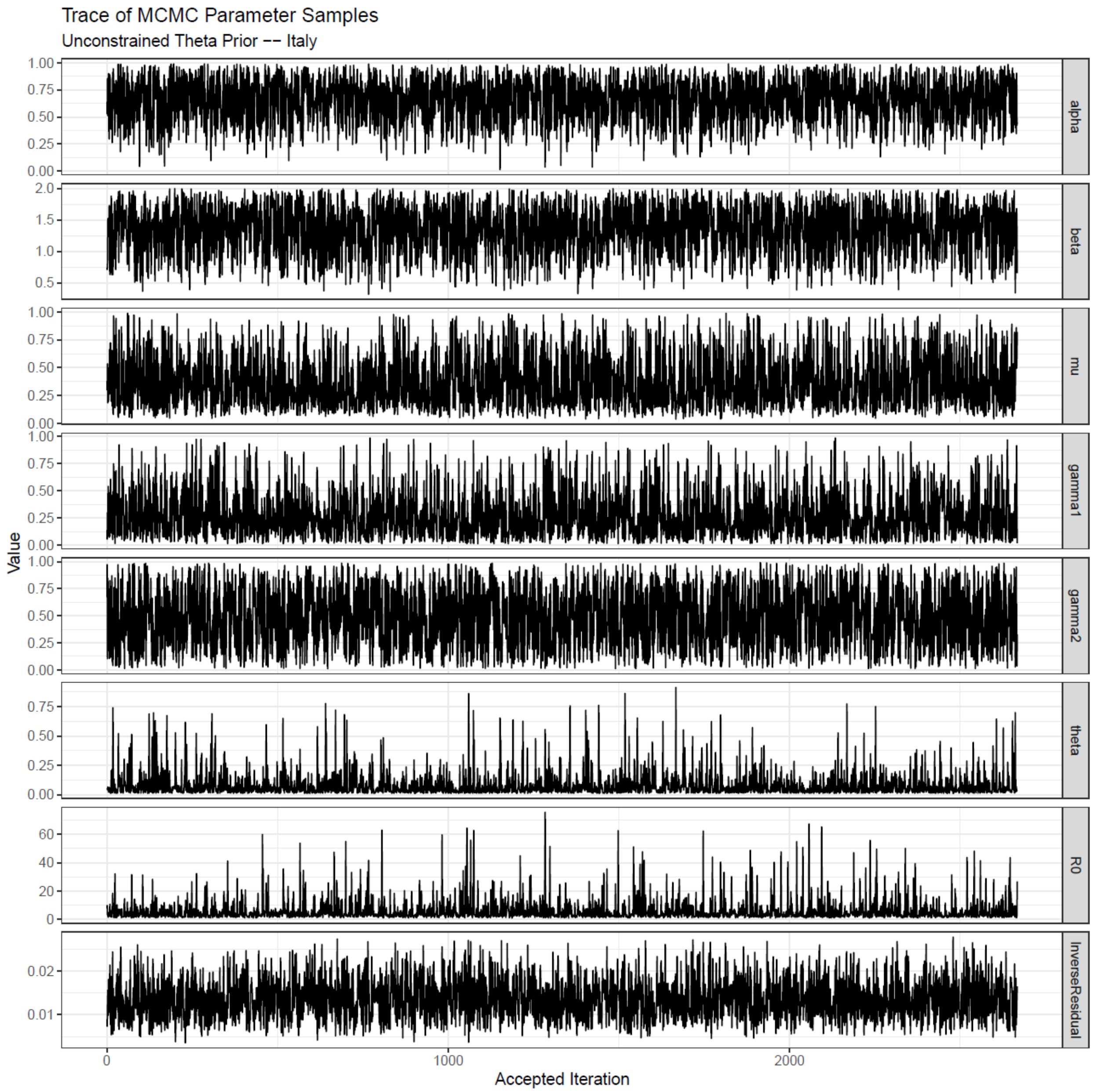

**Figure S16.** The trace plot of accepted parameters using the Metropolis-Hasting algorithm after thinning and burn-in are shown for Italy with unconstrained priors of θ.

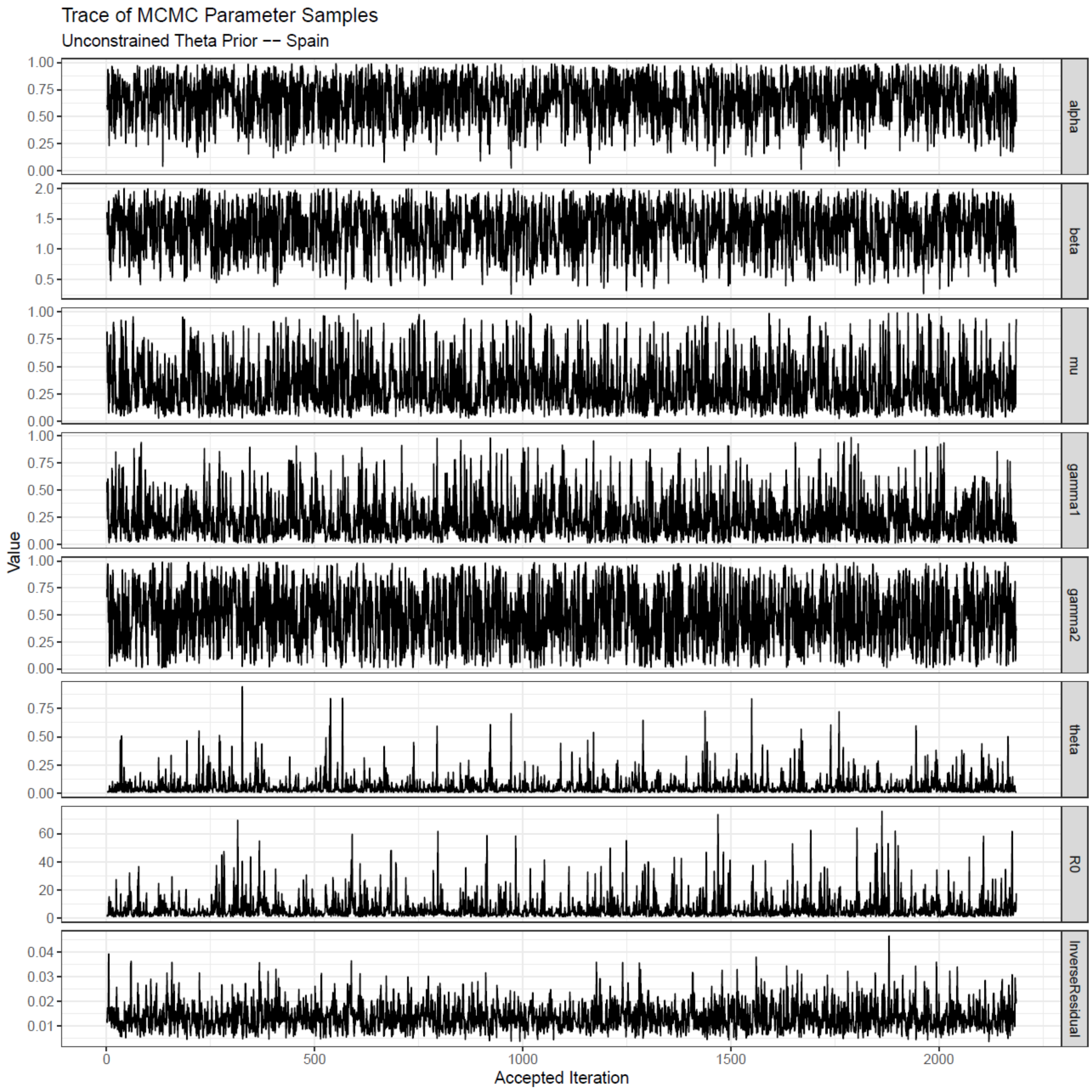

**Figure S17.** The trace plot of accepted parameters using the Metropolis-Hasting algorithm after thinning and burn-in are shown for Spain with unconstrained priors of θ.

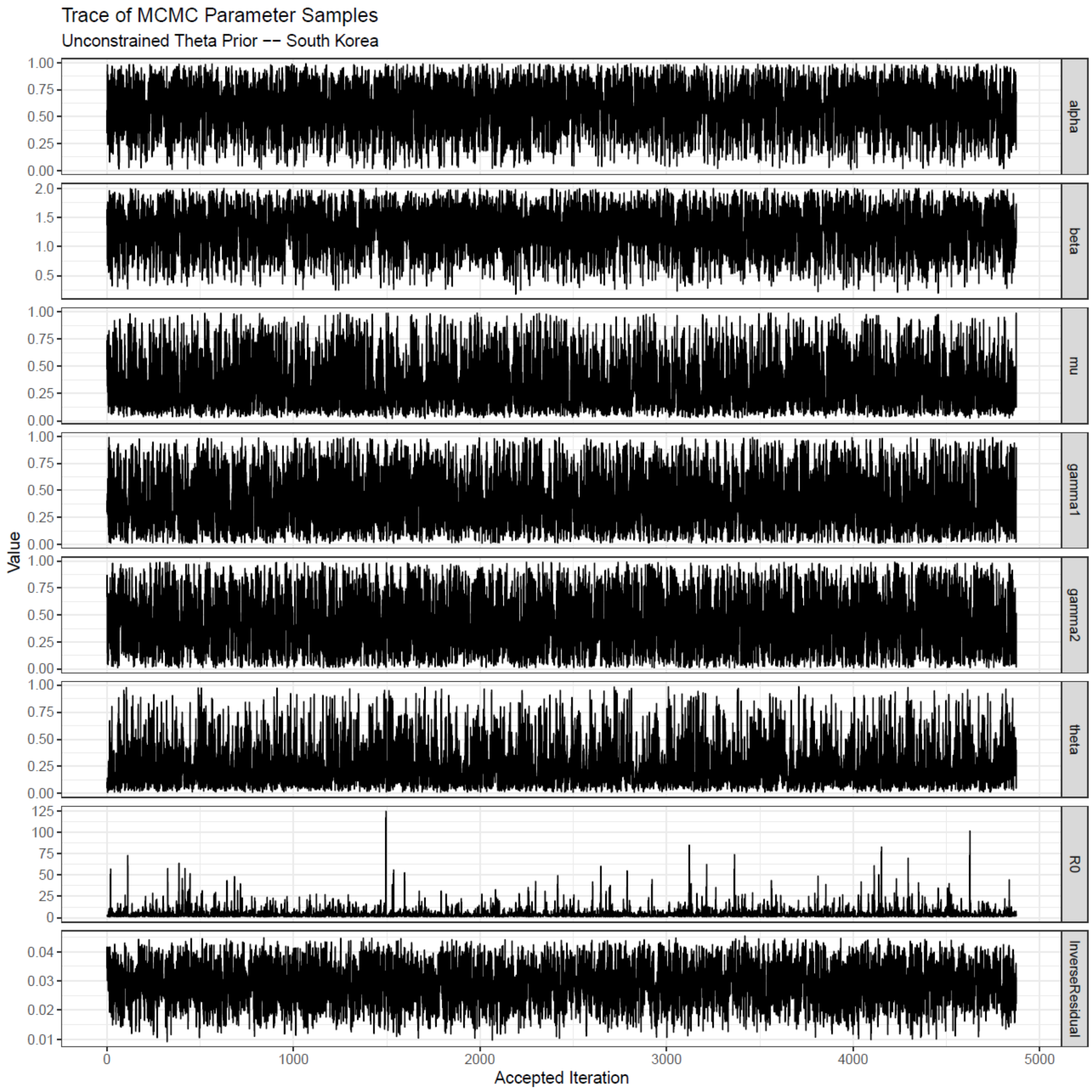

**Figure S18.** The trace plot of accepted parameters using the Metropolis-Hasting algorithm after thinning and burn-in are shown for South Korea with unconstrained priors of θ.

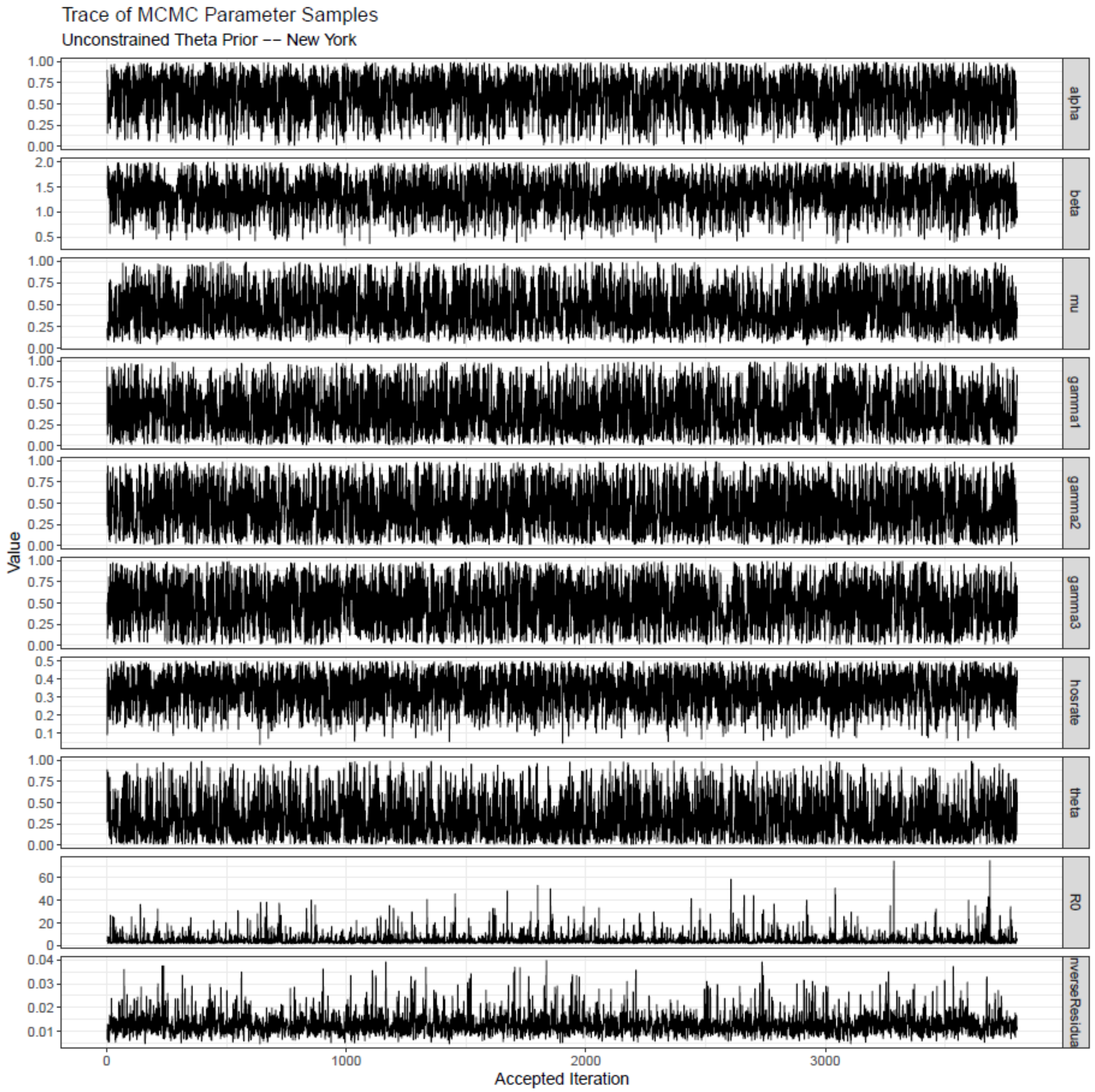

**Figure S19.** The trace plot of accepted parameters using the Metropolis-Hasting algorithm after thinning and burn-in are shown for New York City with unconstrained priors of θ.

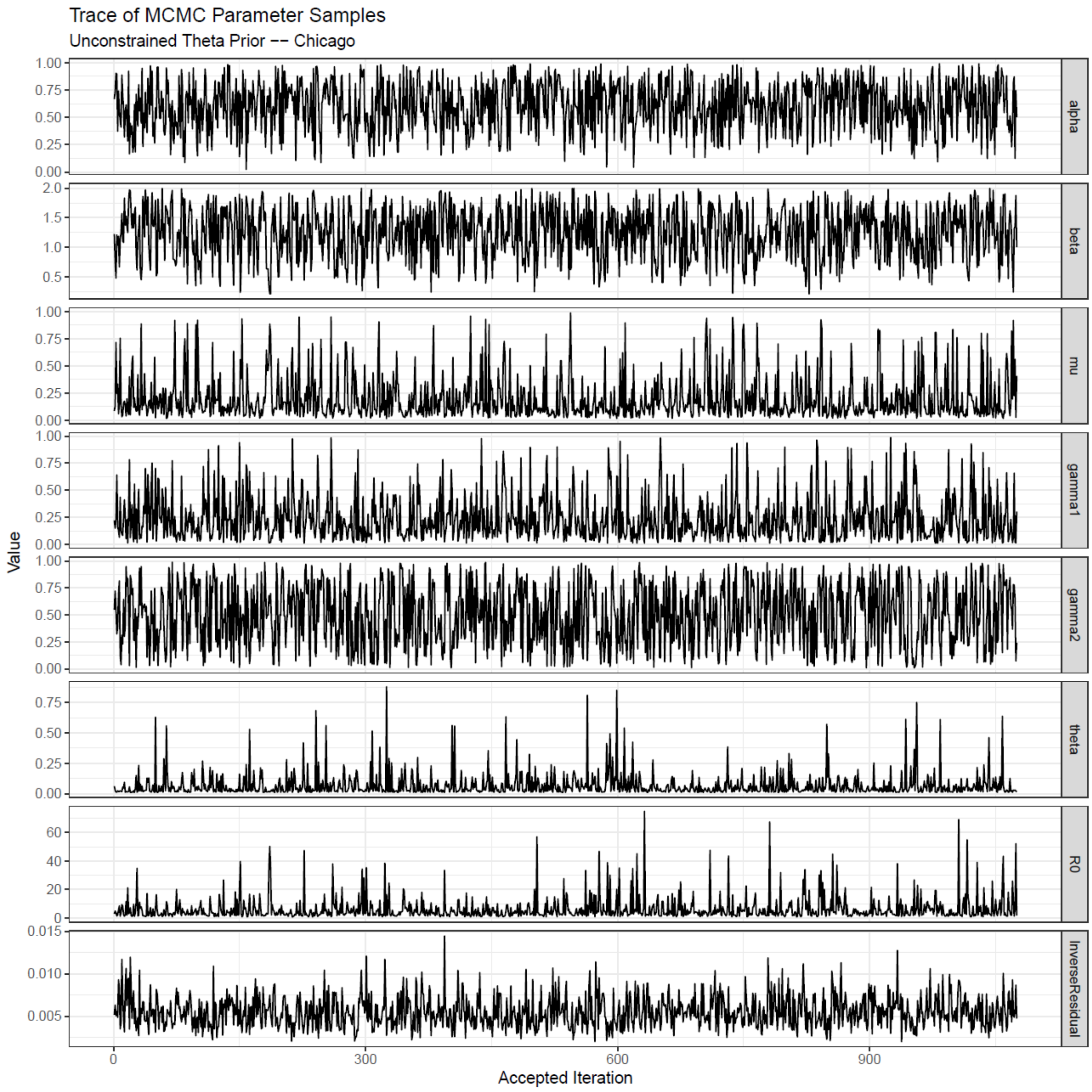

**Figure S20.** The trace plot of accepted parameters using the Metropolis-Hasting algorithm after thinning and burn-in are shown for Chicago with unconstrained priors of θ.

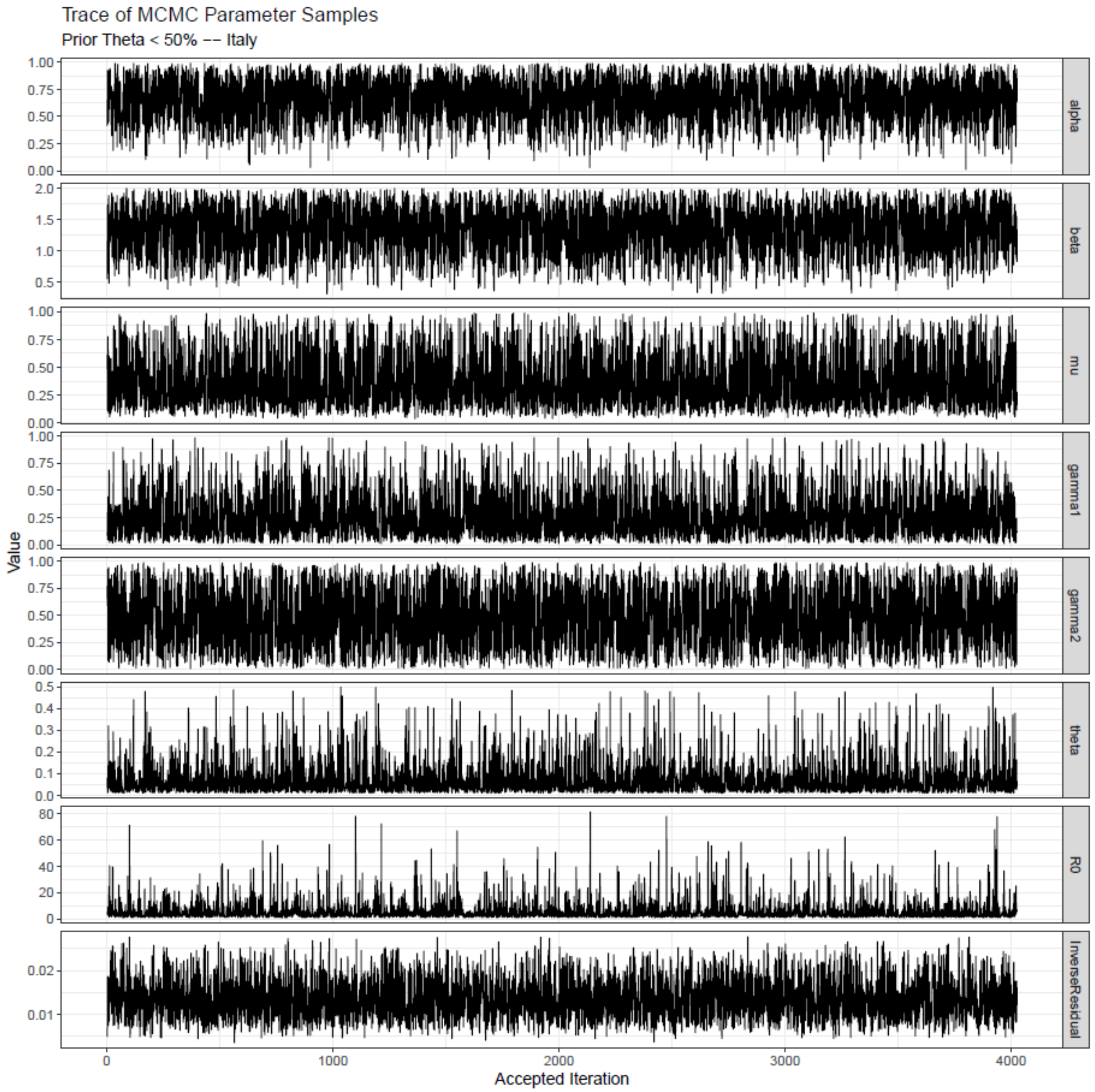

**Figure S21.** The trace plot of accepted parameters using the Metropolis-Hasting algorithm after thinning and burn-in are shown for Italy with priors of θ < 50%.

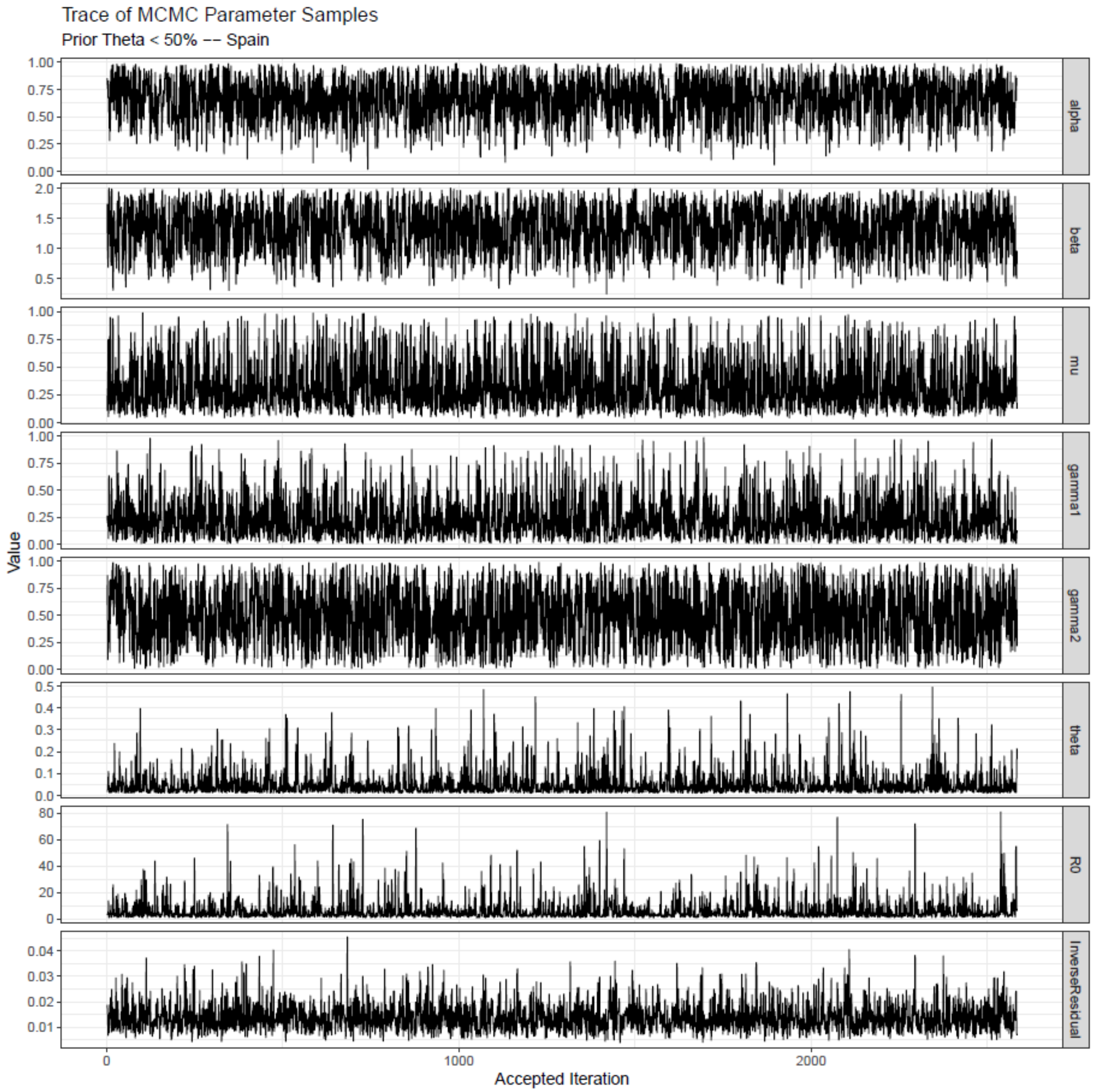

**Figure S22.** The trace plot of accepted parameters using the Metropolis-Hasting algorithm after thinning and burn-in are shown for Spain with priors of θ < 50%.

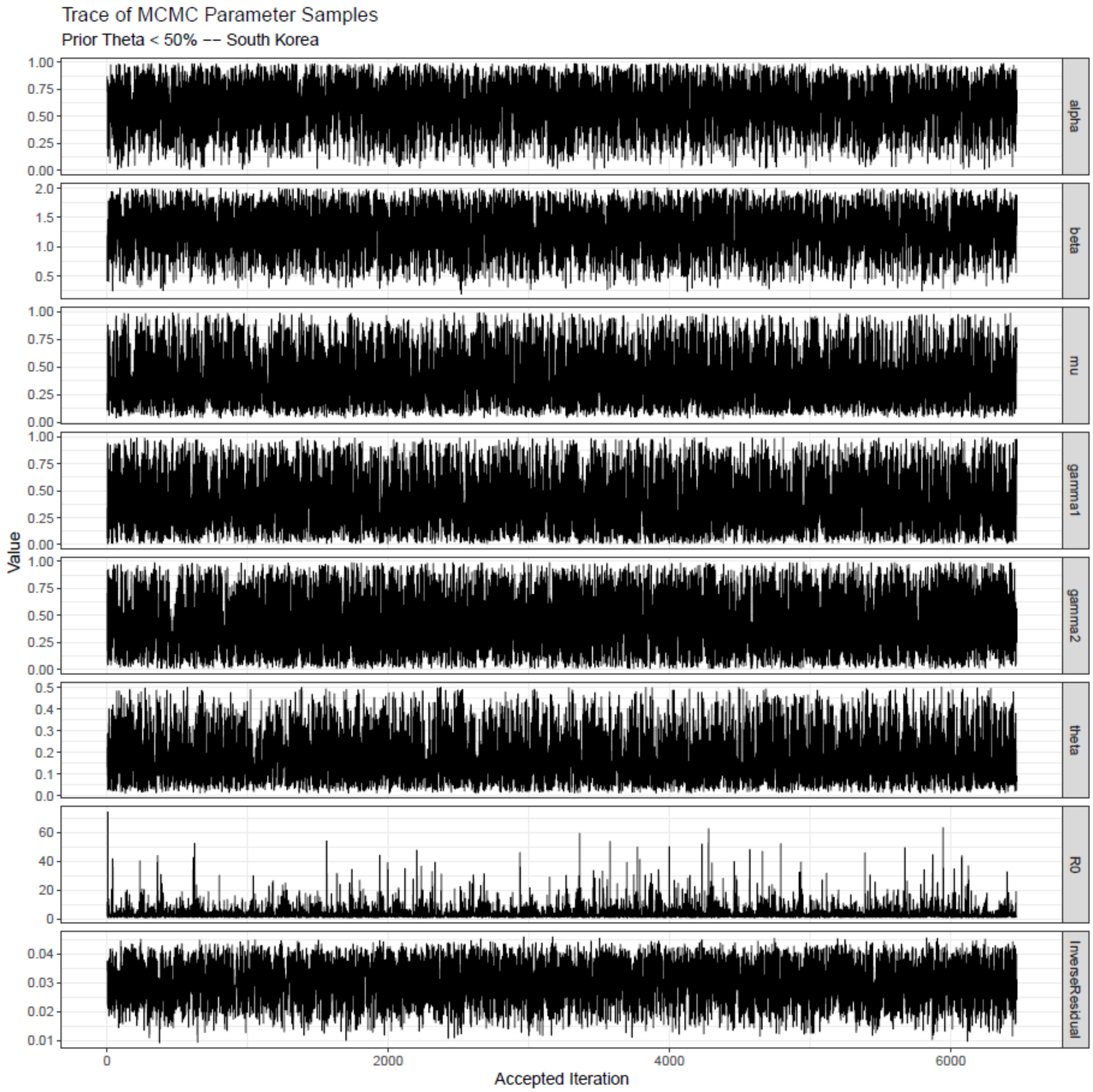

**Figure S23.** The trace plot of accepted parameters using the Metropolis-Hasting algorithm after thinning and burn-in are shown for South Korea with priors of θ < 50%.

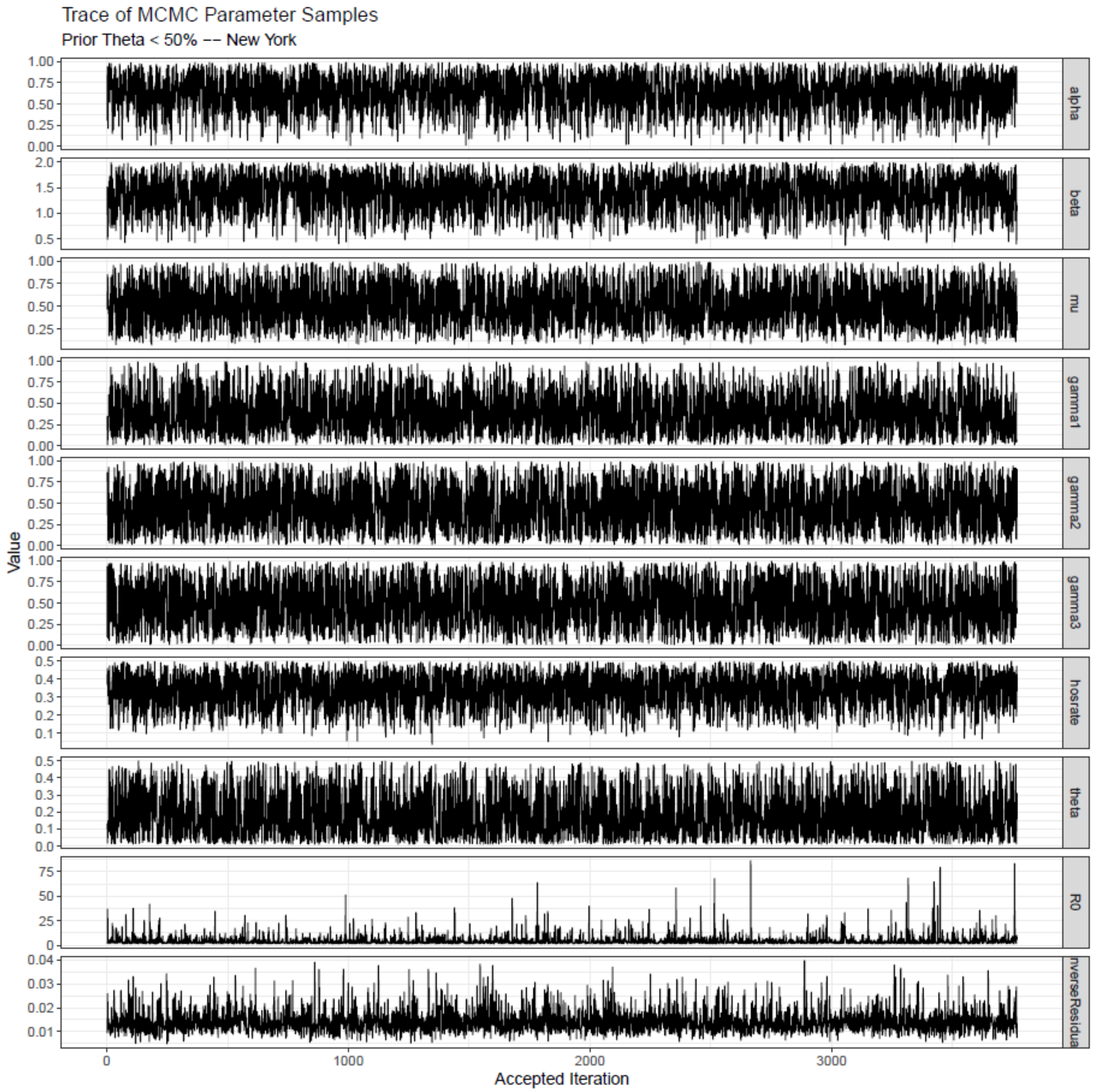

**Figure S24.** The trace plot of accepted parameters using the Metropolis-Hasting algorithm after thinning and burn-in are shown for New York City with priors of θ < 50%.

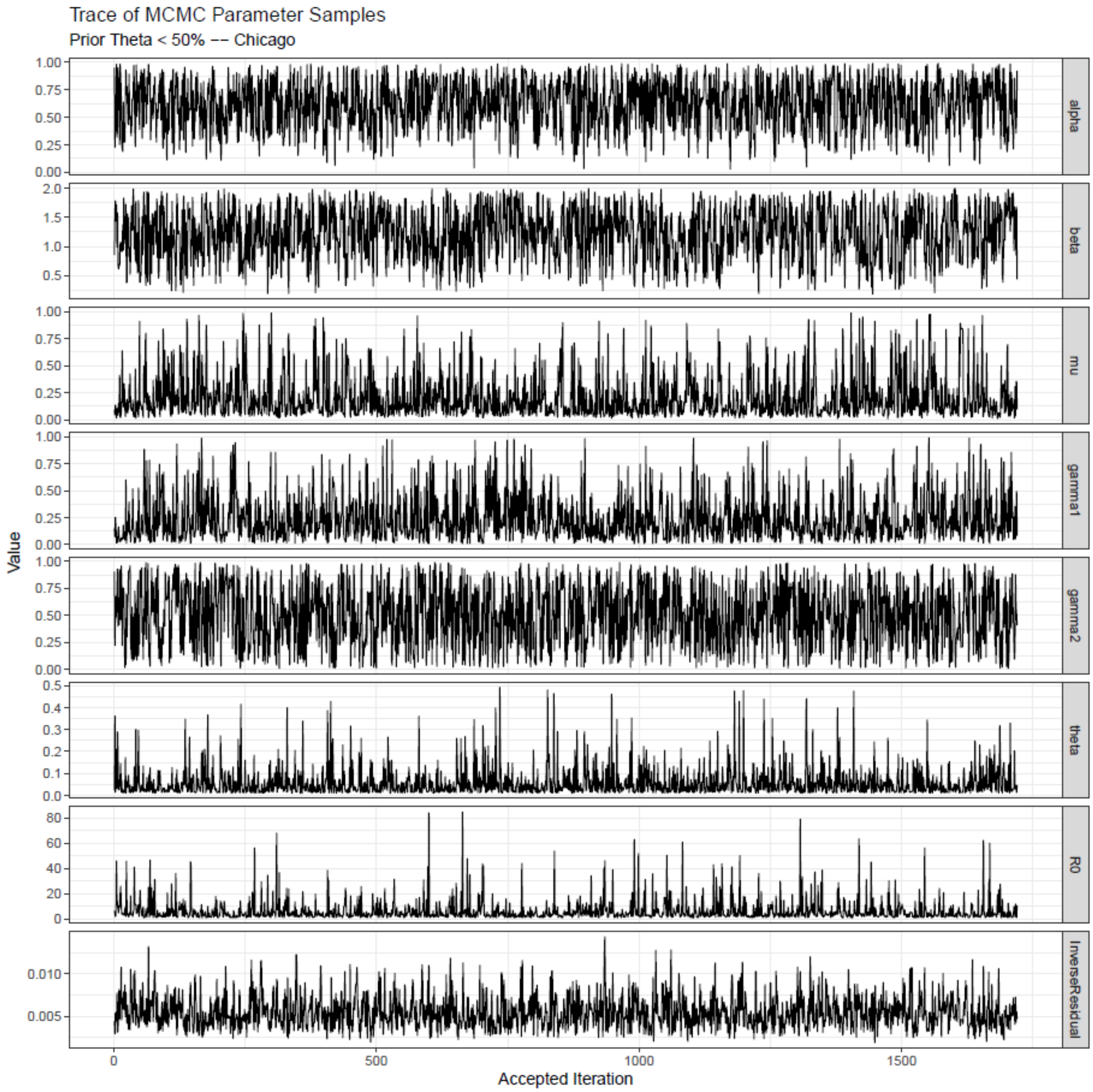

**Figure S25.** The trace plot of accepted parameters using the Metropolis-Hasting algorithm after thinning and burn-in are shown for Chicago with priors of $\theta < 50\%$.

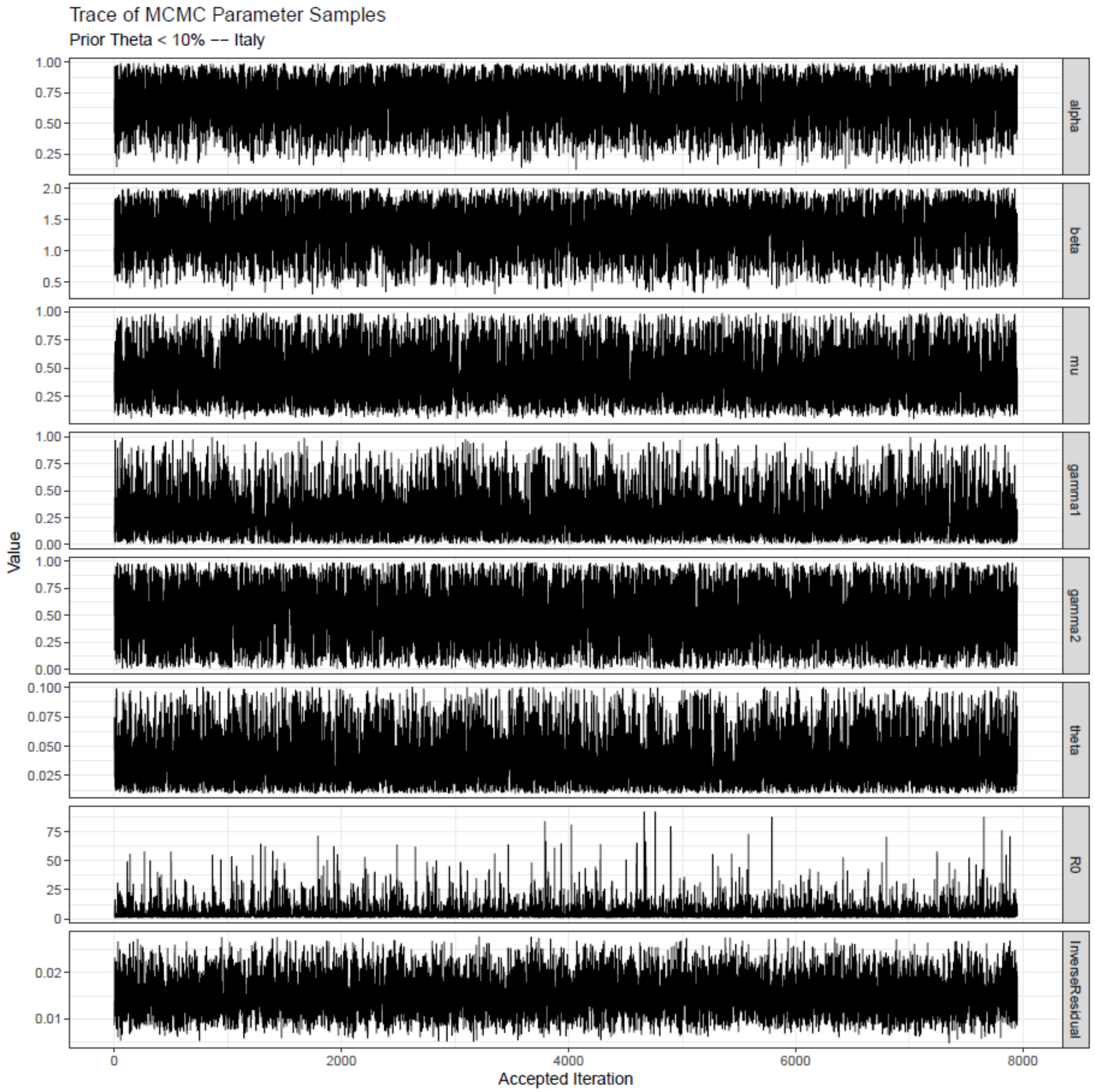

**Figure S26.** The trace plot of accepted parameters using the Metropolis-Hasting algorithm after thinning and burn-in are shown for Italy with priors of $\theta < 10\%$.

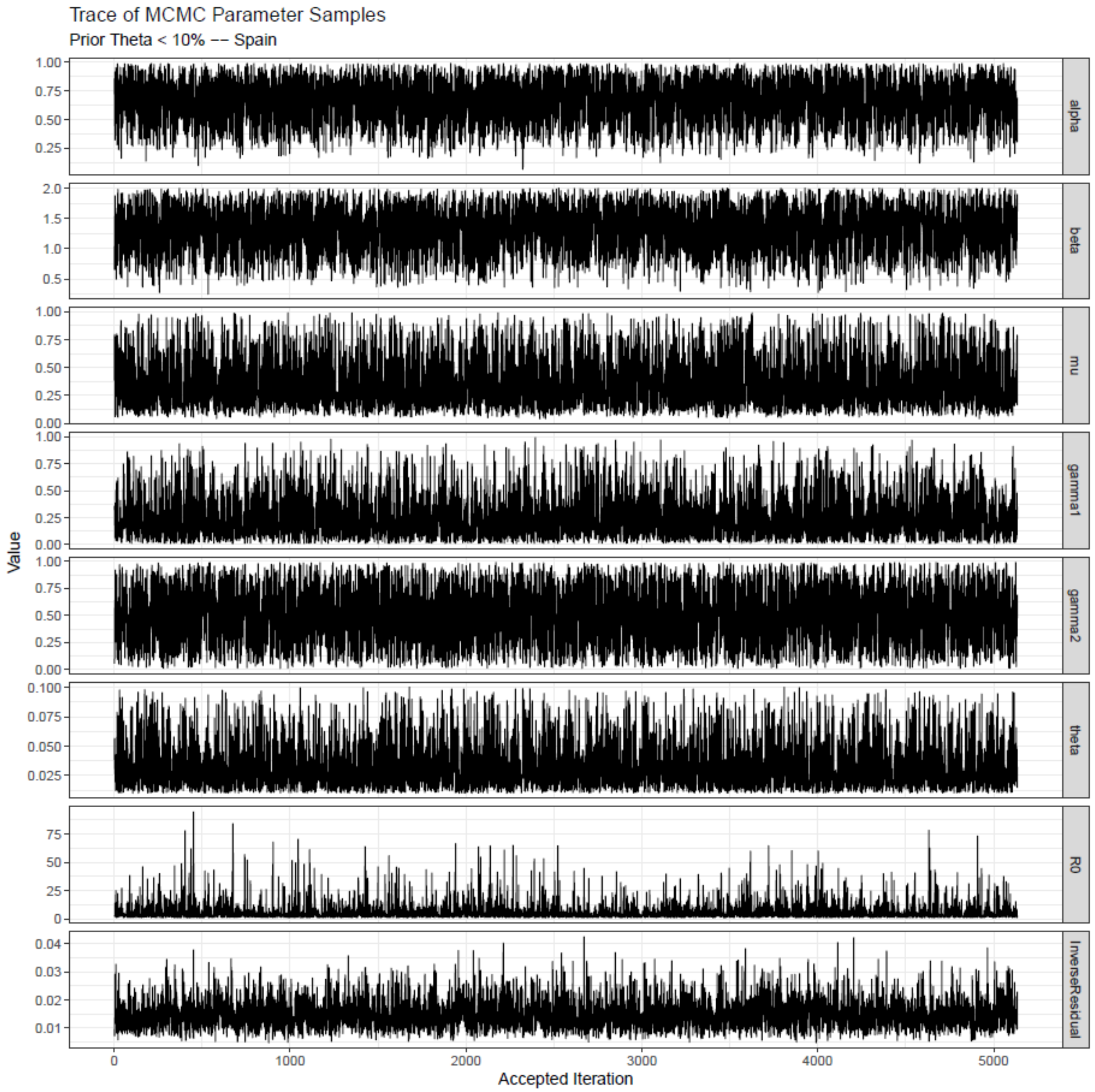

**Figure S27.** The trace plot of accepted parameters using the Metropolis-Hasting algorithm after thinning and burn-in are shown for Spain with priors of θ < 10%.

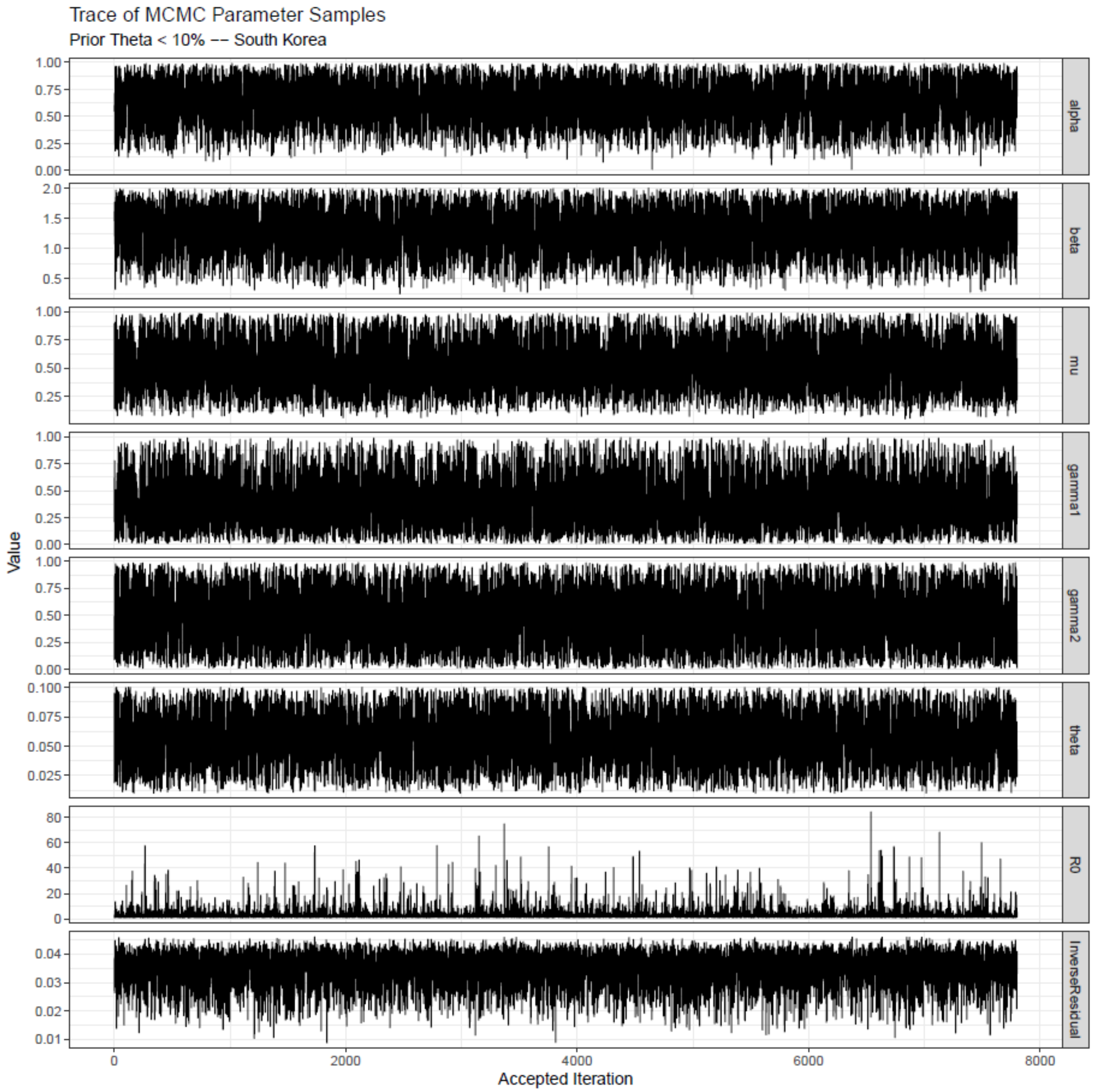

**Figure S28.** The trace plot of accepted parameters using the Metropolis-Hasting algorithm after thinning and burn-in are shown for South Korea with priors of θ < 10%.

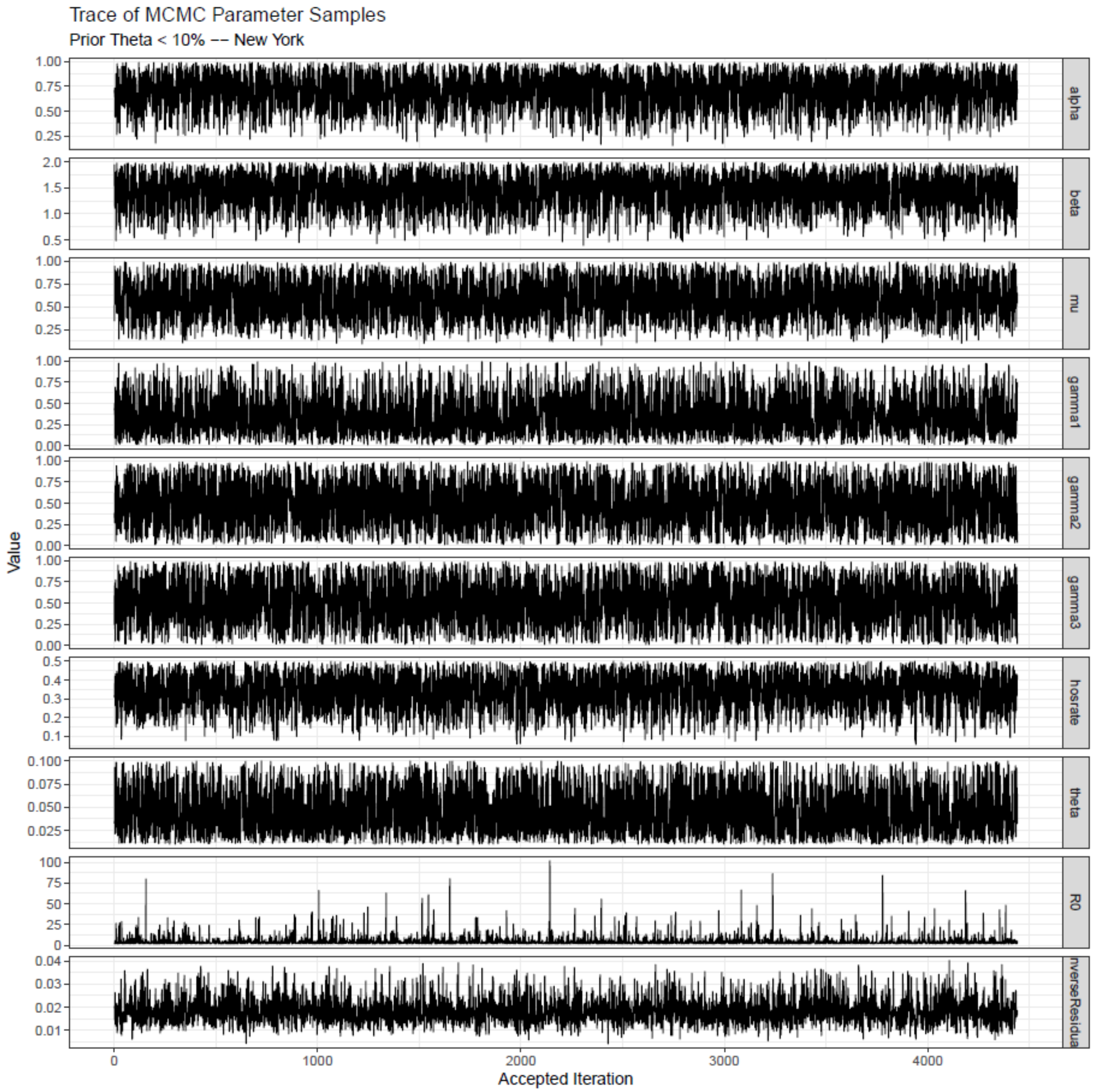

**Figure S29.** The trace plot of accepted parameters using the Metropolis-Hasting algorithm after thinning and burn-in are shown for New York City with priors of θ < 10%.

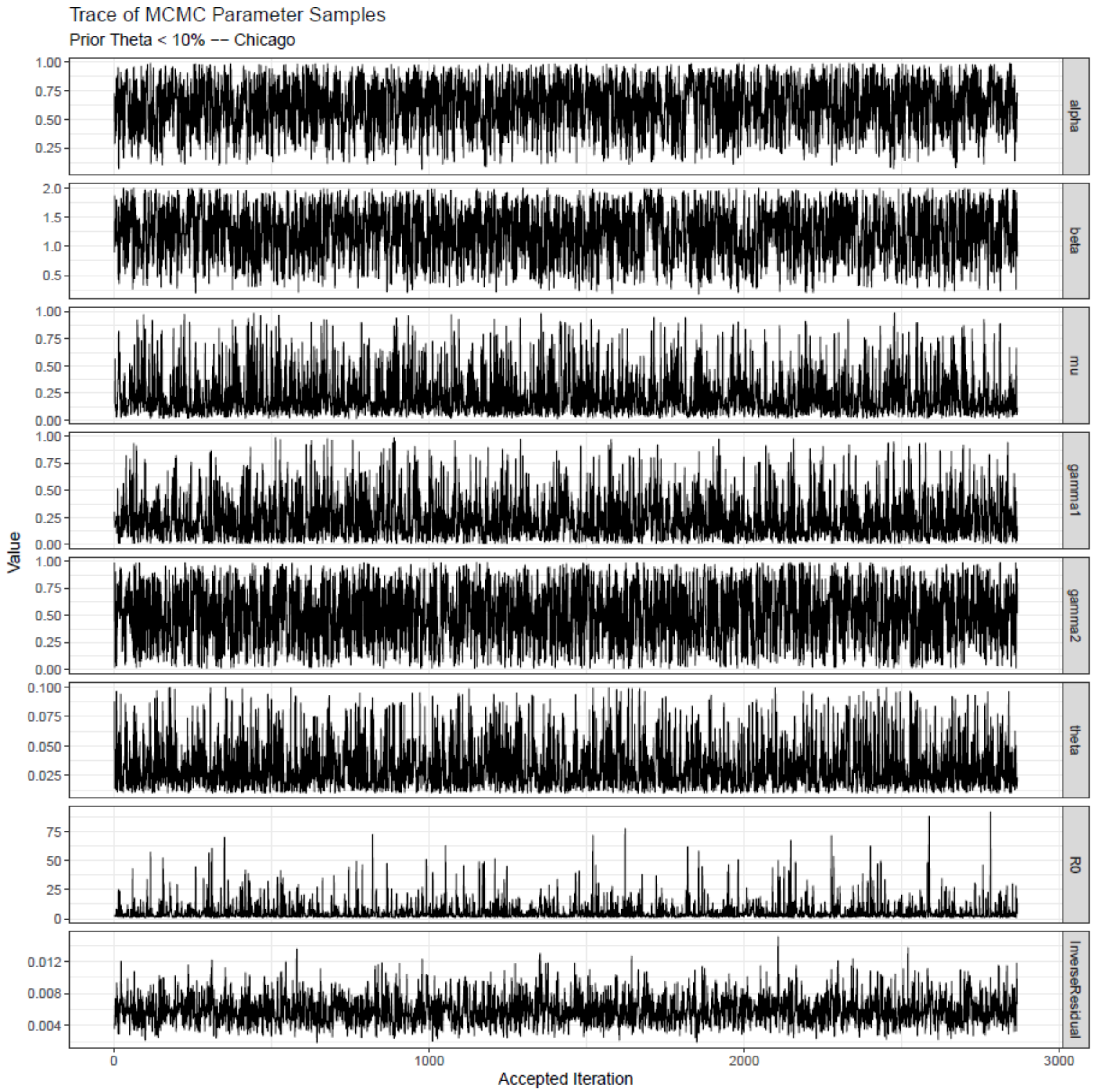

**Figure S30.** The trace plot of accepted parameters using the Metropolis-Hasting algorithm after thinning and burn-in are shown for Chicago with priors of θ < 10%.

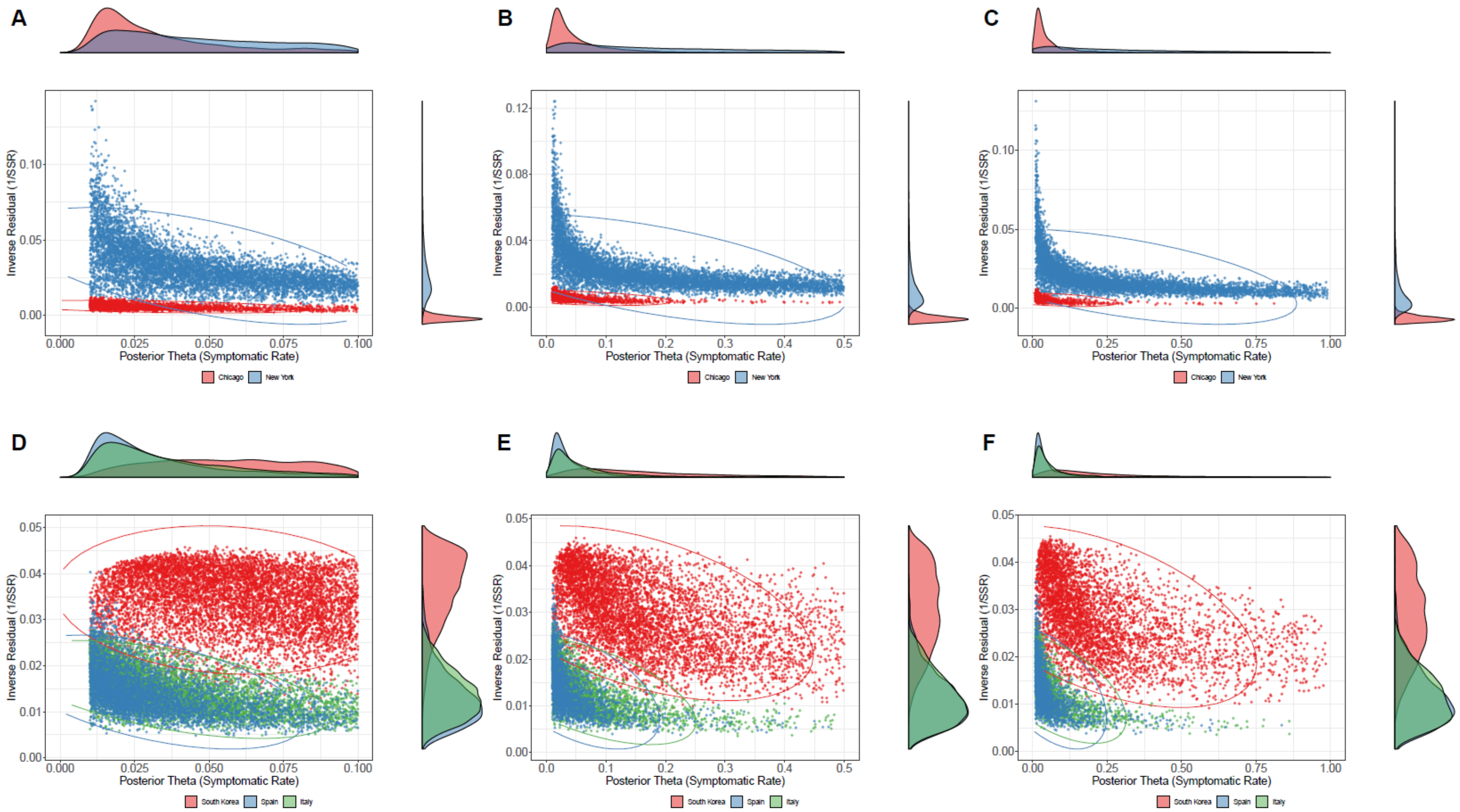

**Figure S31.** Using Markov Chain Monte Carlo (MCMC) methods, we sampled from the posterior distribution of the model parameters for three Bayesian priors which assumes $\theta$ is less than 10%, 50% and 99%. The posterior distribution of $\theta$ and the inverse of sum of squared residuals ($SSR^{-1}$) for Chicago and New York City are shown in A, B, and C, and the same set of priors for Italy, Spain, and South Korea are shown in D, E, and F. The marginal posterior density of θ and $SSR^{-1}$ are displayed on the right and top of the scatter plot, respectively. The ellipses encircle 95% of simulation runs for each location.

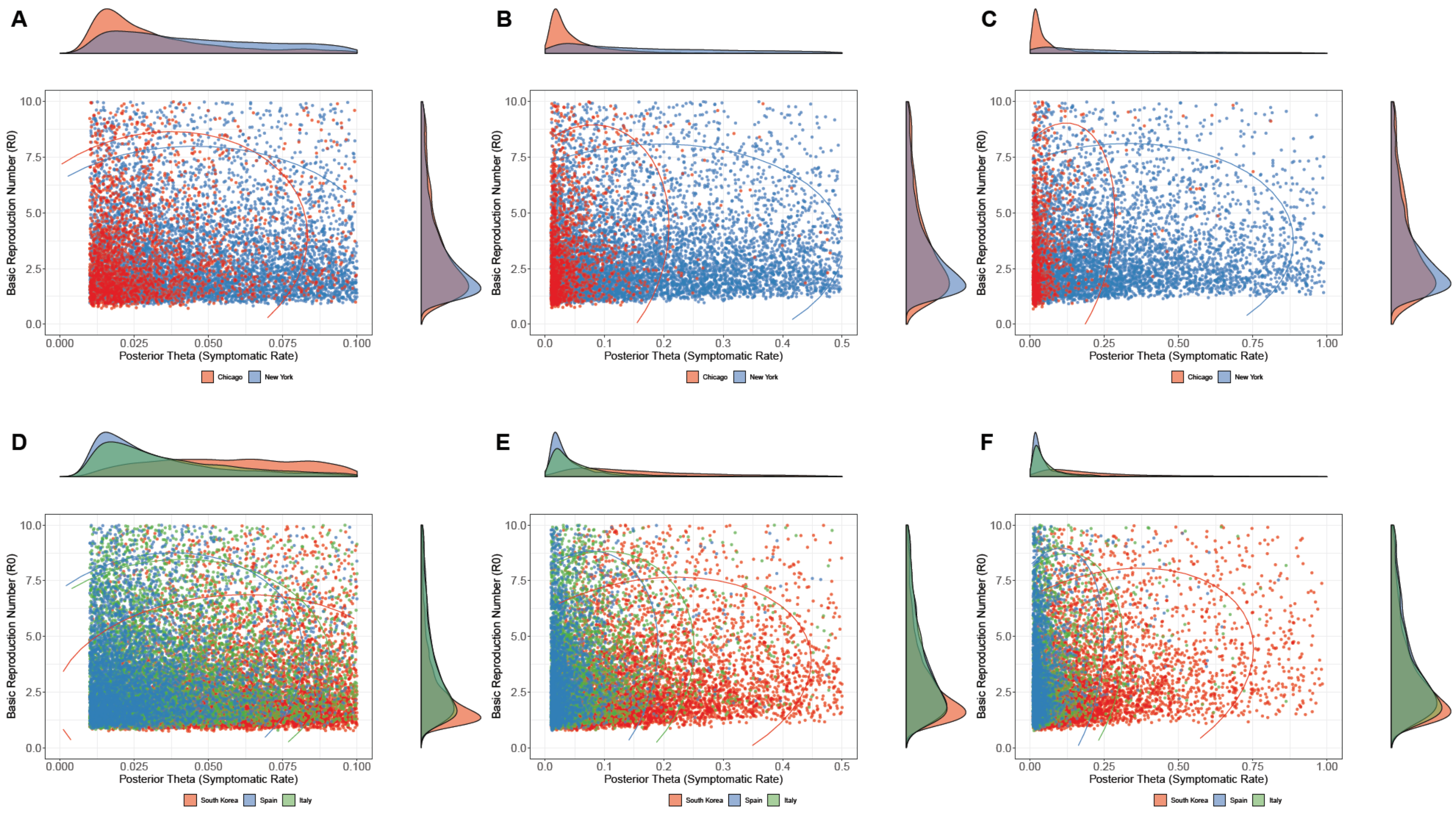

**Figure S32.** Using Markov Chain Monte Carlo (MCMC) methods, we sampled from the posterior distribution of the model parameters for three Bayesian priors which assumes $\theta$ is less than 10%, 50% and 99%. The posterior distribution of $\theta$ and $R_0$ for the three priors of Chicago and New York City are shown in A, B, and C, and the same set of priors for Italy, Spain, and South Korea are shown in D, E, and F. The marginal posterior density of $R_0$ and $\theta$ are displayed on the right and top of the scatter plot, respectively. The ellipses encircle 95% of simulation runs for each location.

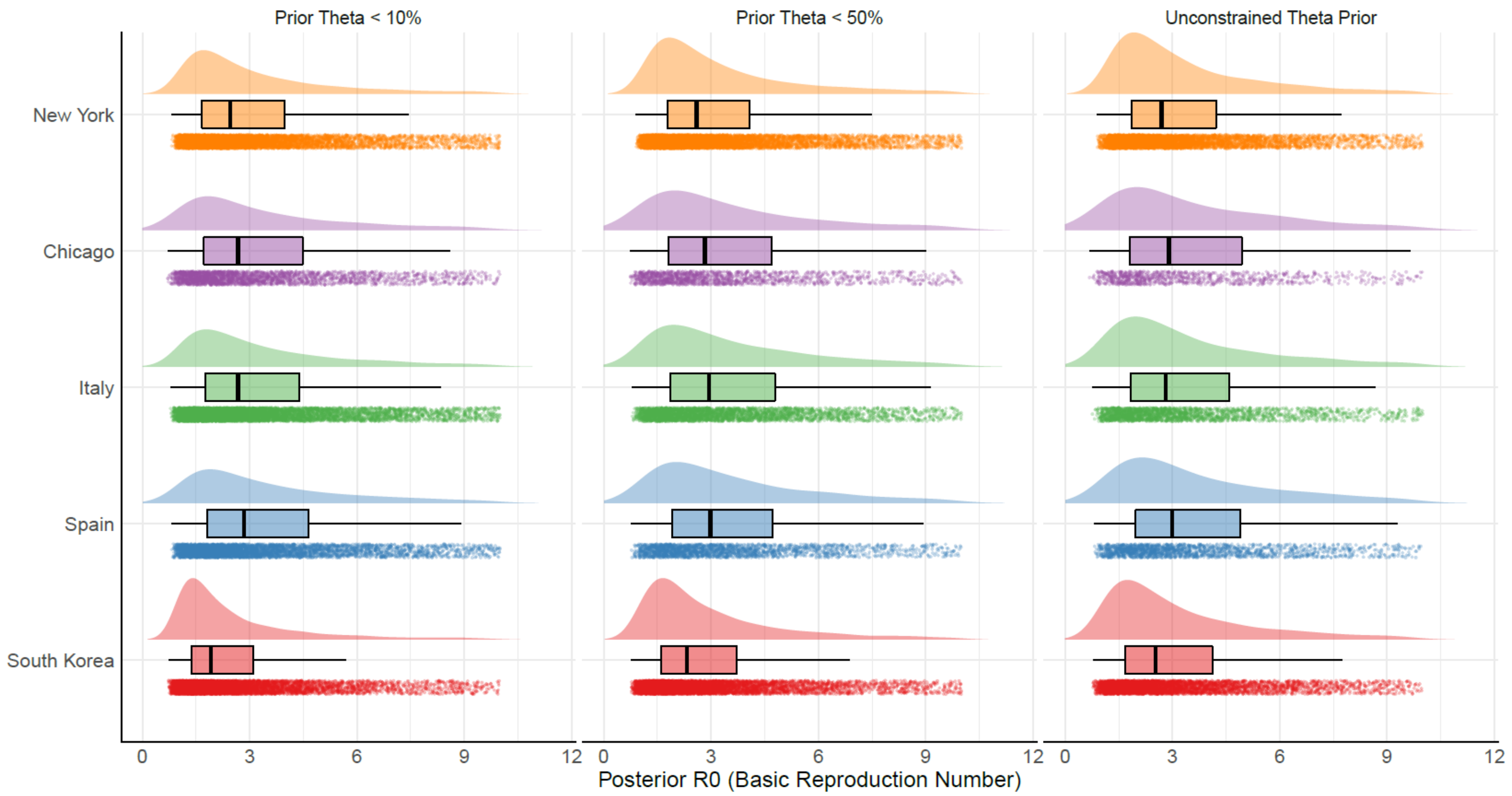

**Figure S33.** The posterior distribution of $R_0$ is generated from the MCMC sampling for each location are shown for the symptomatic rate $\theta$. The three groupings represent the Bayesian priors for $\theta$, where θ<10%, θ <50%, and $\theta$ is unconstrained.

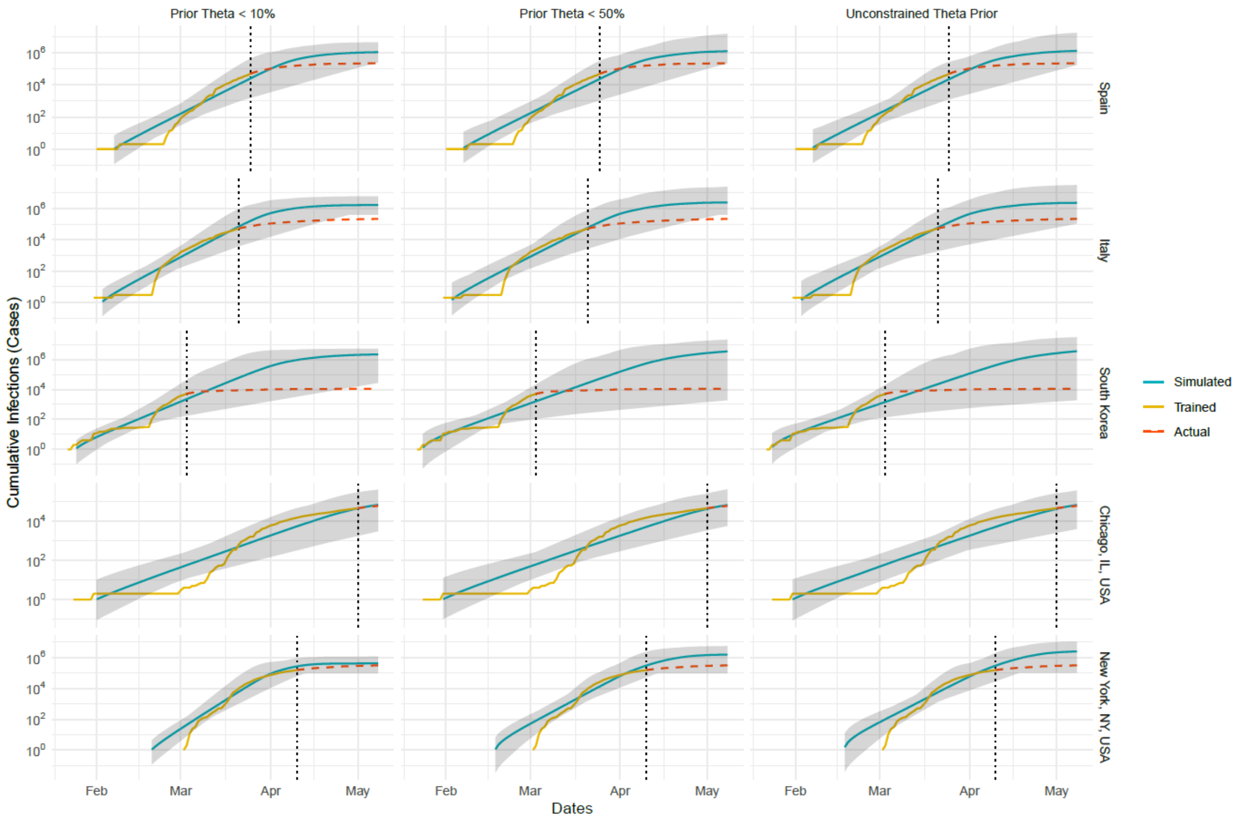

**Figure S34.** The simulated and actual cumulative infections for each location (Spain, Italy, South Korea, Chicago, IL, and New York City, NY are shown) for three different prior distributions of the symptomatic rate ($\theta$). The turquoise line (simulated) corresponds with the simulated outputs for cumulative confirmed cases, which were fitted to the yellow line (trained) representing the actual data. The red dashed line represents the actual data after the explosion date, which is demarcated by the vertical black dotted line. The grey ribbon represents the upper and lower bounds of the MCMC sampling for all cases for each day.

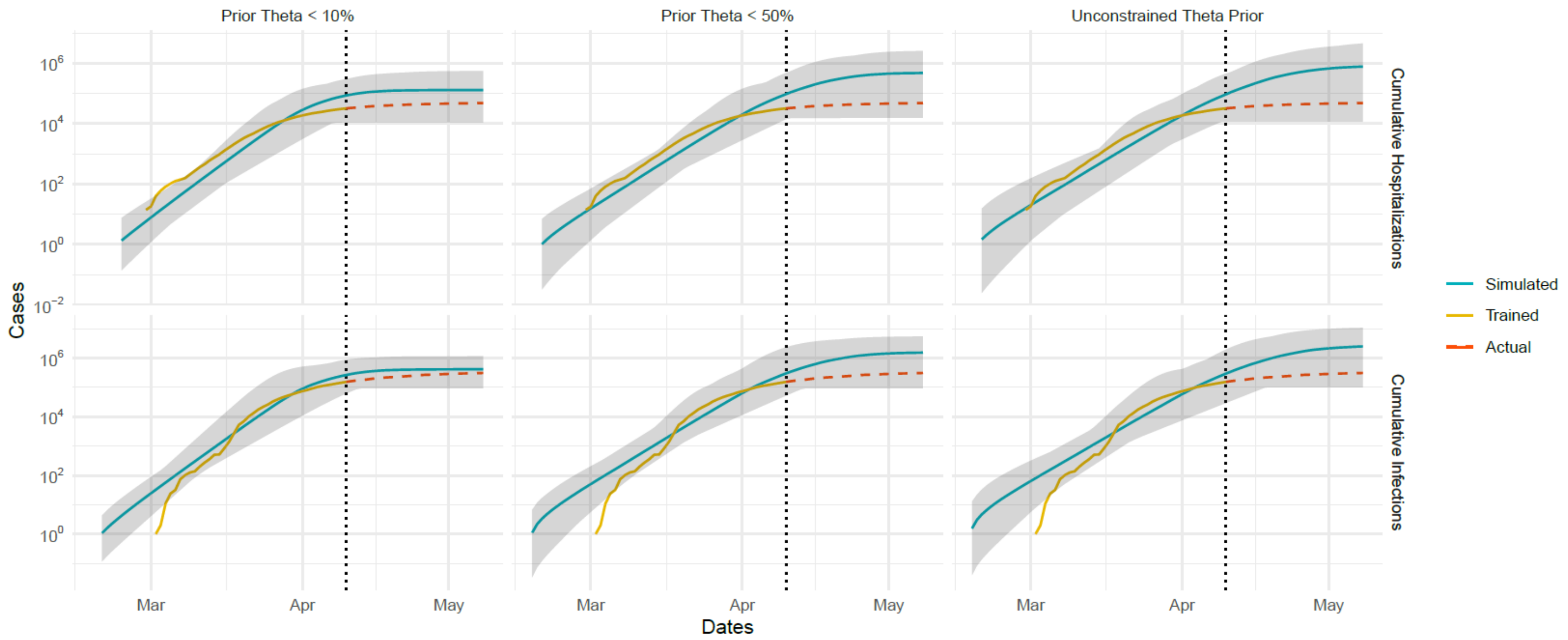

**Figure S35.** Simulated and actual cumulative hospitalizations and cumulative infections for New York City for three different prior distributions of the symptomatic rate ($\theta$). The turquoise line (simulated) corresponds with the simulated outputs for cumulative confirmed cases, which were fitted to the yellow line (trained) representing the actual data. The red dashed line represents the actual data after the explosion date which is demarcated by the vertical black dotted line. The grey ribbon represents the upper and lower bounds of the MCMC sampling for all cases for each day.
